# Transparent and robust Artificial intelligence-driven Electrocardiogram model for Left Ventricular Systolic Dysfunction

**DOI:** 10.1101/2024.10.06.24314872

**Authors:** Min Sung Lee, Jong-Hwan Jang, Sora Kang, Ga In Han, Ah-Hyun Yoo, Yong-Yeon Jo, Jeong Min Son, Joon-myoung Kwon, Sooyeon Lee, Ji Sung Lee, Hak Seung Lee, Kyung-Hee Kim

## Abstract

Heart failure (HF) is an escalating global health concern, worsened by an aging population and limitations in traditional diagnostic methods like electrocardiograms (ECG). The advent of deep learning has shown promise for utilizing 12-lead ECG models for the early detection of left ventricular systolic dysfunction (LVSD), a crucial HF indicator. This study validates the AiTiALVSD, an AI/machine learning-enabled Software as a Medical Device, for its effectiveness, transparency, and robustness in detecting LVSD. Conducted at Mediplex Sejong Hospital in the Republic of Korea, this retrospective single-center cohort study involved patients suspected of LVSD. The AiTiALVSD model, which is based on a deep learning algorithm, was assessed against echocardiography findings. To improve model transparency, the study utilized Testing with Concept Activation Vectors (TCAV) and included clustering analysis and robustness tests against ECG noise and lead reversals. The study involved 688 participants and found AiTiALVSD to have a high diagnostic performance, with an AUROC of 0.919. There was a significant correlation between AiTiALVSD scores and left ventricular ejection fraction values, confirming the model’s predictive accuracy. TCAV analysis showed the model’s alignment with medical knowledge, establishing its clinical plausibility. Despite its robustness to ECG artifacts, there was a noted decrease in specificity in the presence of ECG noise. AiTiALVSD’s high diagnostic accuracy, transparency, and resilience to common ECG discrepancies underscore its potential for early LVSD detection in clinical settings. This study highlights the importance of transparency and robustness in AI/ML-based diagnostics, setting a new benchmark in cardiac care.

## Introduction

Heart failure (HF) shows a global prevalence of 1-2% primarily due to an aging population.^1,2^ With HF associated with poor short-term and long-term survival rates, early screening and diagnosis have become increasingly critical.^3^ Echocardiography stands as the gold standard for HF diagnosis, assessing ejection fraction (EF) for risk stratification. Yet, its resource-demanding nature has spurred the search for a more cost-effective and accessible diagnostic method, which remains an unmet need.^4^ Recent advancements include deep learning-based 12-lead electrocardiogram (ECG) models for left ventricular systolic dysfunction (LVSD) screening, sparking extensive research.^5,6^ Our previous work introduced a high-accuracy deep learning-based 12-lead ECG model for LVSD detection (AiTiALVSD).^7–10^

The medical artificial intelligence (AI) domain is swiftly advancing, with growing demands for not just performance but also transparency and robustness.^11,12^ This shift acknowledges the intricacies of diagnostics, highlighting the need for healthcare professionals to navigate AI applications in medicine with confidence. Despite notable advances in medical image analysis, AI-ECG analysis research lags, especially in aspects of transparency.^13,14^ Despite notable advances in medical image analysis, AI-ECG analysis research lags, especially in aspects of transparency.^15^ Addressing this, we have developed a dedicated dataset and embarked on research focusing on reliability and clinical applicability. Our approach includes comprehensive and quantitative evaluations of AI models for ECG analysis, aiming to bridge the current research gap.^11,13,16,17,18^

## Results

### Study population

Between March 2017 and August 2021, 7,834 patients at Mediplex Sejong Hospital, undergoing echocardiography and ECG, were screened, selecting 688 for the clinical trial’s analysis set; seven were excluded due to ECG data quality, resulting in 681 participants. The cohort’s mean age was 60.7 ± 14.8 years, with males representing 51%. Detailed demographics and clinical profiles are in Table 1. Analysis highlighted older age and higher rates of diabetes, coronary artery disease, and chronic kidney disease in the LVSD group compared to the non-LVSD group. Data collection flow is illustrated in Figure 2.

**Table 1.** Baseline characteristics.

|  | LVSD<br>n=98 | Non-LVSD<br>n=583 | p value |
| --- | --- | --- | --- |
| Age | $67.5 \pm 13.6$ | $59.6 \pm 14.7$ | <0.001 |
| Male | 67 (68.4) | 283 (48.5) | <0.001 |
| Height | $164.7 \pm 9.3$ | $163.6 \pm 9.3$ | 0.299 |
| Weight | $66.8 \pm 13.8$ | $66.8 \pm 13.4$ | 0.963 |
| Medical History |  |  |  |
| Hypertension | 48 (49.0) | 240 (41.2) | 0.147 |
| Diabetes mellitus | 48 (49.0) | 93 (16.0) | <0.001 |
| Coronary artery disease | 36 (36.7) | 68 (11.7) | <0.001 |
| Chronic kidney disease | 14 (14.3) | 17 (2.9) | <0.001 |
| LVEF, % | $33.2 \pm 6.3$ | $64.0 \pm 6.9$ | <0.001 |
| Echo-ECG time difference, hours | $1.5 \pm 3.6$ | $3.0 \pm 5.9$ | 0.02 |
| AiTIALVSD score | $46.3 \pm 28.2$ | $2.6 \pm 6.6$ | <0.001 |

Values are expressed as n (%), mean ± standard deviation.
LVEF, left ventricular ejection fraction; Echo, echocardiography; ECG, electrocardiography; LVSD, left ventricular systolic dysfunction

**Figure 1.**
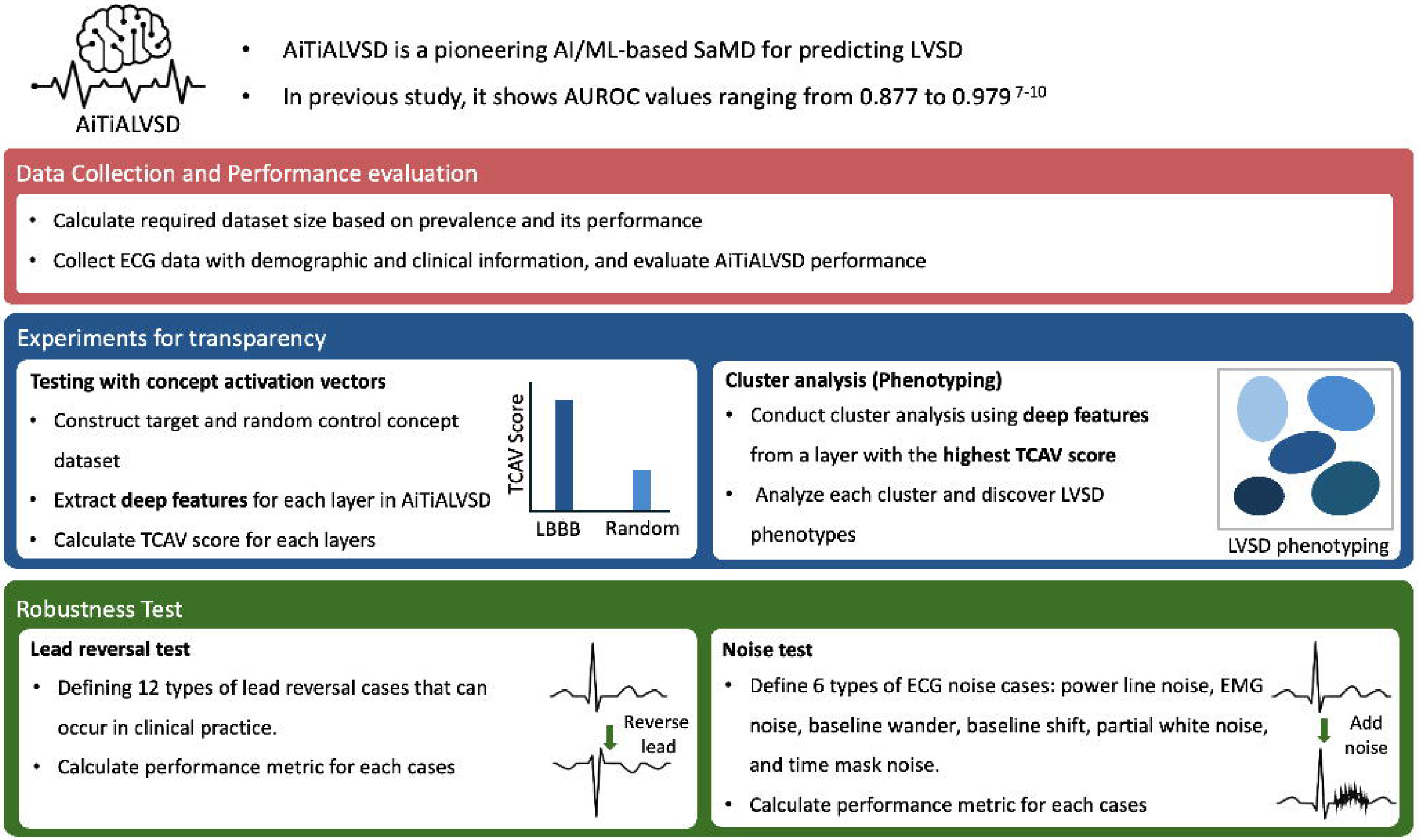
Study flow for evaluation of AiTiALVSD. It outlines the methodology for validating the AiTiALVSD, a pretrained model for predicting Left Ventricular Systolic Dysfunction (LVSD). Initially, data collection protocols are established to gather a statistically significant sample and relevant demographic and clinical data. The model’s performance is then evaluated by selecting appropriate metrics and comparison targets. Transparency is addressed through experiments involving concept activation vectors (TCAV) and cluster analysis, which are used to understand model decisions and discover LVSD phenotypes. Finally, robustness tests are conducted, including a reverse test to assess performance under various lead reversal cases, and a noise test to evaluate the model’s resilience to different types of ECG noise. This structured process aims to ensure the model’s performance, transparency, and robustness for application in real-world. LBBB, Left bundle branch block; Electrocardiogram, ECG; TCAV, Testing with concept activation vectors; EMG, electromyography.

**Figure 2.**
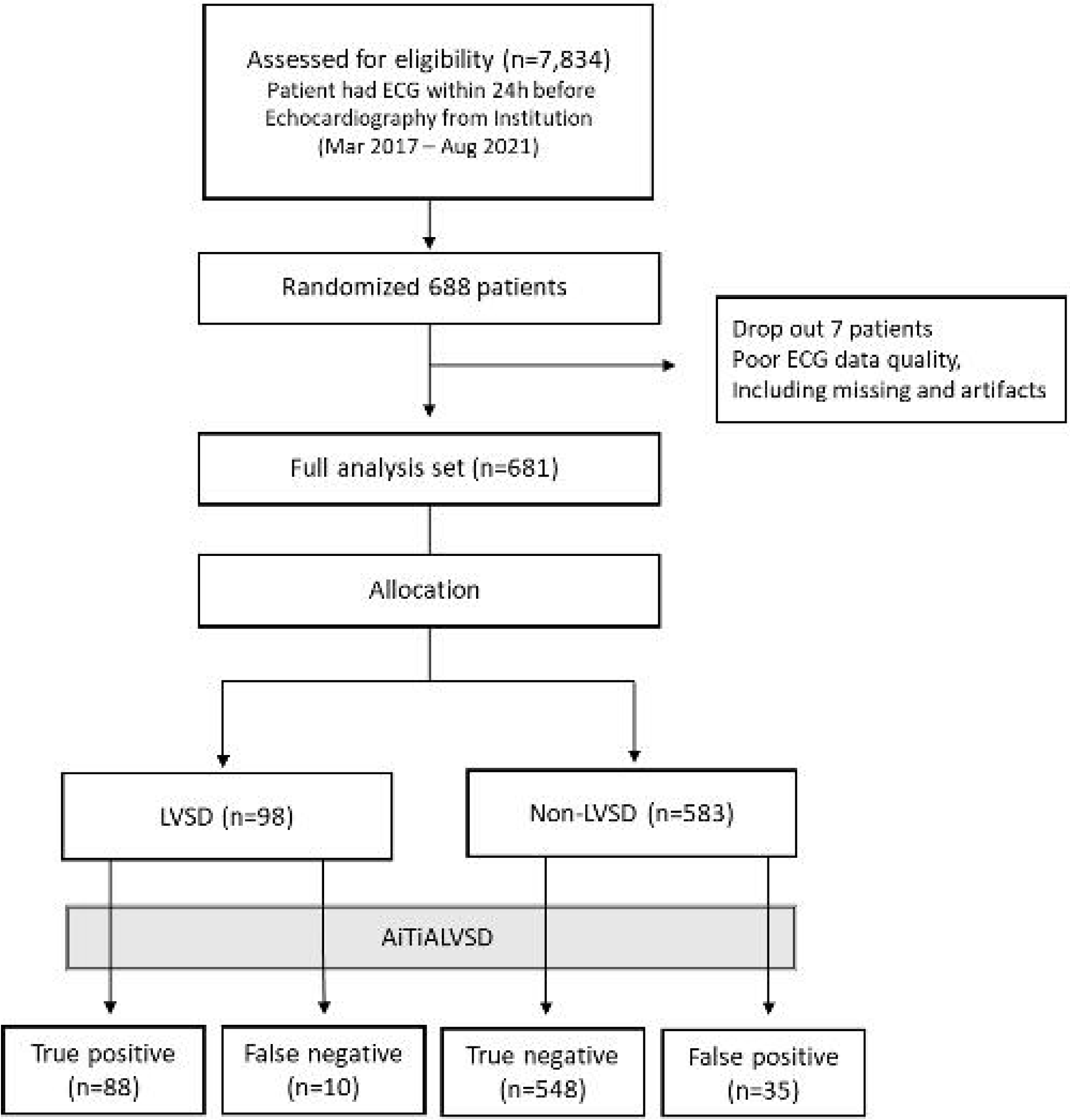
Data flow. During the study period, among 7,834 ECG-echocardiography pairs that met the inclusion and exclusion criteria, 688 were selected for the eligibility; the noise assessment module filtered out 7 ECG-echocardiography pairs, and the final data of 681 pairs (98 LVSD, 593 Non-LVSD) were selected as the full analysis set.

### AI-ECG model performance

AiTiALVSD demonstrated a diagnostic performance for LVSD with an AUROC of 0.919 (95% CI 0.887-0.951), with its score distribution detailed in Supplementary Figure 2. The LVSD group’s scores predominantly clustered below 5.0, peaking at 53.5. Notably, AiTiALVSD scores inversely correlated with LVEF values, evidenced by a Pearson R of - 0.781 (p<0.001), as shown in Supplementary Figure 3.

Performance metrics outlined in Table 2 reveal AUROC, sensitivity, specificity, PPV, and NPV scores of 0.971, 0.898, 0.940, 0.715, and 0.982, respectively. Performance differentiation for EF thresholds of 35% and 50% showed AUROCs of 0.906 (95% CI 0.869-0.943) and 0.891 (95% CI 0.857-0.925), respectively, detailed in Supplementary Table 1.

**Table 2.** AiTiALVSD classification performance.

|  | <sup>a</sup>AUROC<br>(95% CI) | <sup>b</sup>AUROC<br>(95% CI) | Sensitivity<br>(95% CI) | Specificity<br>(95% CI) | PPV<br>(95% CI) | NPV<br>(95% CI) |
| --- | --- | --- | --- | --- | --- | --- |
| AiTIALVSD | 0.919<br>(0.887-0.951) | 0.971<br>(0.954-0.989) | 0.898<br>(0.838-0.958) | 0.940<br>(0.921-0.959) | 0.715<br>(0.636-0.795) | 0.982<br>(0.971-0.993) |

<sup>a</sup> AUROC with binary output
<sup>b</sup> AUROC with continuous output
PPV, positive predictive value; NPV, negative predictive value; CI, confidence interval

Subgroup analyses, accounting for variations in clinical characteristics and ECG patterns, maintained robustness with AUROCs above 0.7, even among those with coronary artery disease, chronic kidney disease, and atrial fibrillation (Supplementary Table 2). Supplementary Table 3 and 4 contrast the performances of NT-proBNP and AiTiALVSD, with AUROCs of 0.72 (95% CI 0.635-0.804) and 0.905 (95% CI 0.842-0.968) among a subset of 96 patients.

### TCAV interpretation

Figure 3 illustrates TCAV’s analysis across AiTiALVSD’s four feature blocks, demonstrating significant engagement with nine LVSD-associated concepts. These concepts, notably Left LBBB, RBBB, AF, and Conduction Disorder, achieved average TCAV scores exceeding 0.8, highlighting their integral role in LVSD prediction. This pattern suggests AiTiALVSD not only accurately identifies these cardiac conditions but also prioritizes their importance in diagnosing LVSD. Particularly, the first feature block showed superior TCAV scores for six concepts (LAD: 0.816, LBBB: 0.984, AF: 0.974, abnormal Q wave: 0.926, abnormal T wave: 0.964, and prolonged QT: 0.922), suggesting a heightened extraction of relevant features for LVSD prediction compared to other blocks. This emphasizes the first block’s extraction of clinically significant features, laying the groundwork for subsequent cluster analysis utilizing these characteristics.

**Figure 3.**
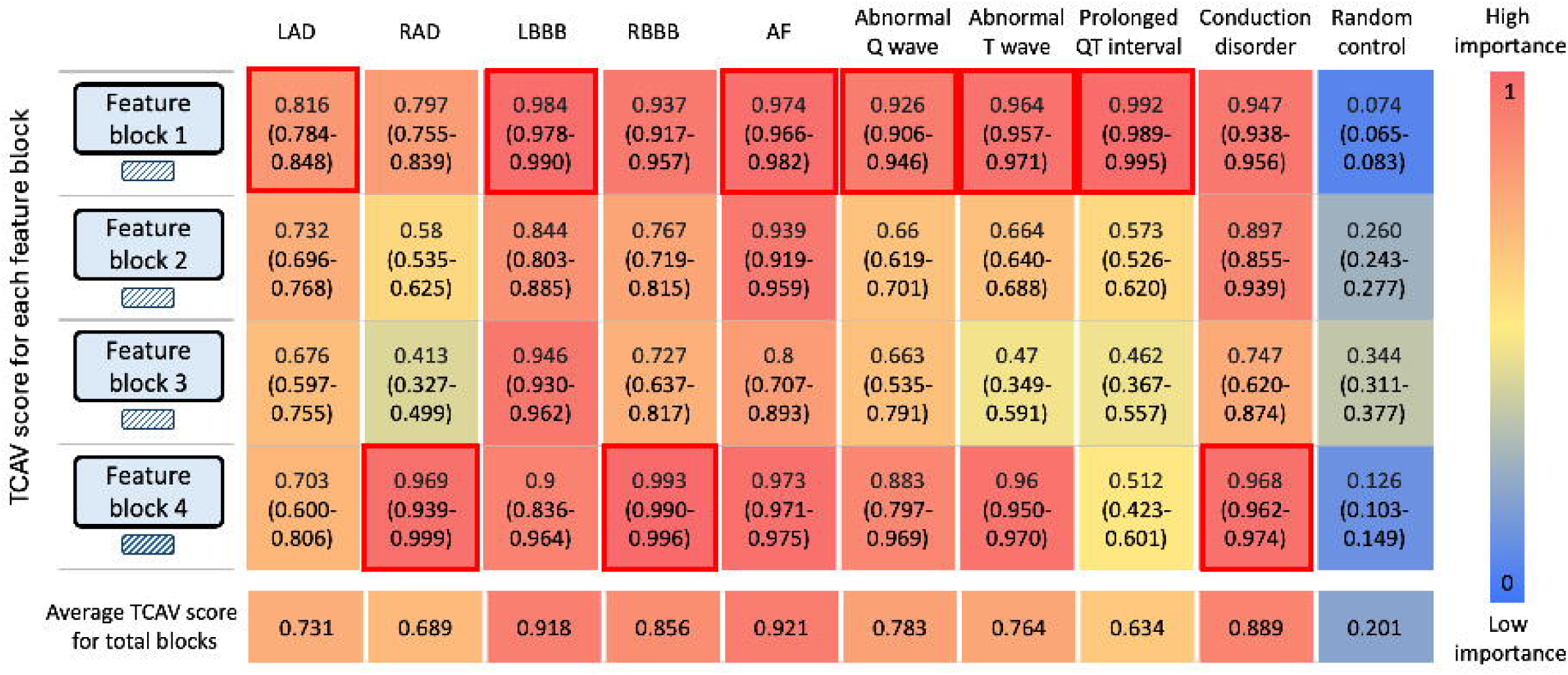
Testing with concept activation vectors of AiTiALVSD. It shows TCAV result extracted from the four feature blocks that make up AiTiALVSD; based on existing medical knowledge, we determined the nine concepts of ECG features corresponding to LVSD high risk and analyzed their impact on LVSD detection by comparing them with a random feature concept that excludes the features; in the first and last blocks, the random features had little importance, but the 9 concept features had high relative importance. LAD, left axis deviation; RAD, right axis deviation; LBBB, left bundle branch block; RBBB, right bundle branch block; AF, atrial fibrillation and atrial flutter.

**Figure 4.**
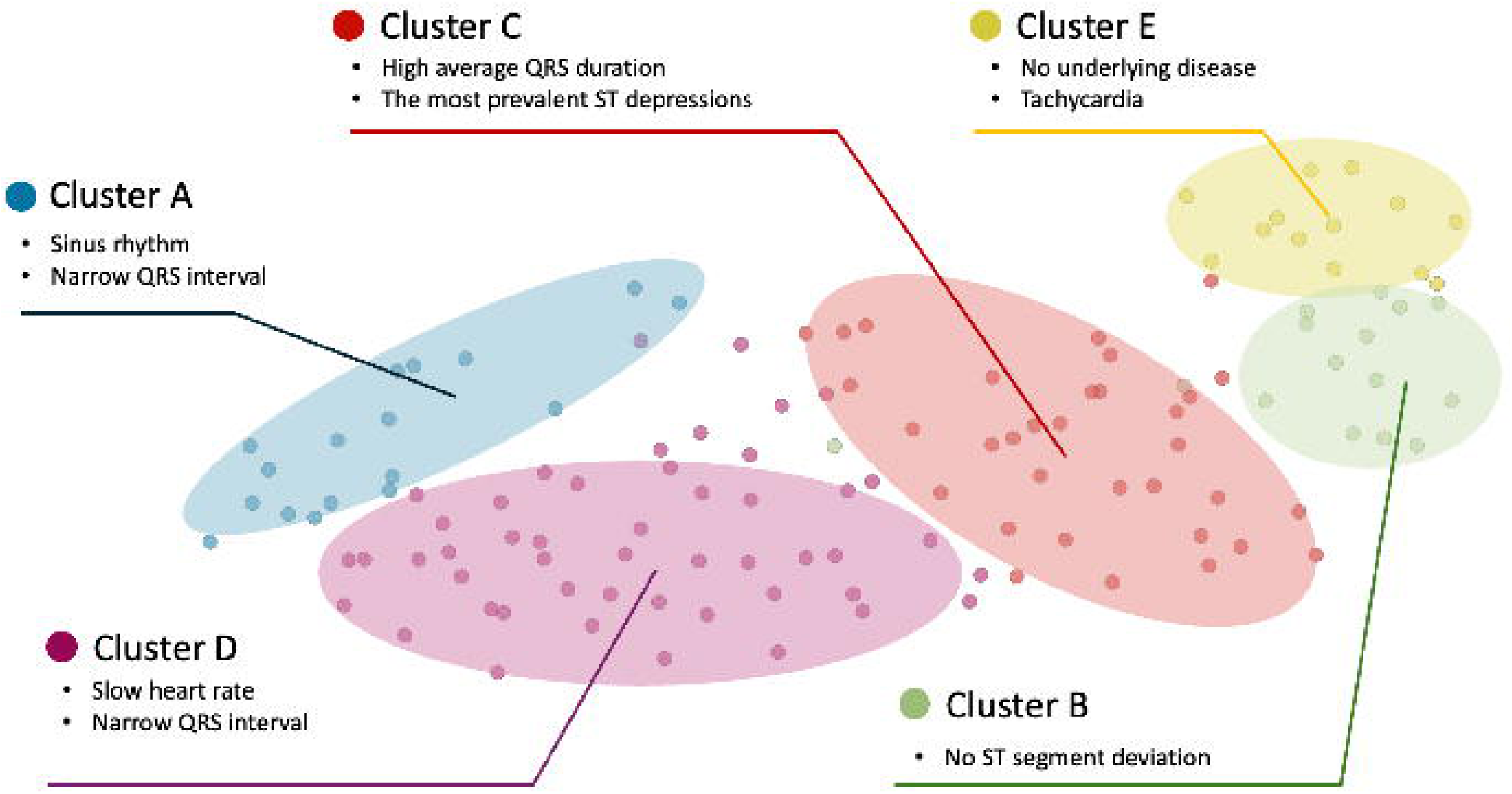
Visualization of clustering analysis using t-distributed Stochastic Neighbor Embedding (T-SNE) This figure presents a two-dimensional scatter plot illustrating five distinct clusters, each representing a different ECG characteristic within a patient cohort. The clusters are differentiated by colors and labeled from A to E, with each cluster denoting unique ECG features.

### Clustering of AiTiALVSD-positive LVSD cases for phenotyping

Leveraging TCAV insights, we hypothesized that the first feature block’s hidden features would closely align with established electrocardiographic knowledge. This premise guided our cluster analysis of LVSD-positive electrocardiograms identified by AiTiALVSD, resulting in five distinct clusters within our development dataset, facilitating a detailed demographic, electrocardiographic, and echocardiographic feature comparison among clusters (Supplementary Text 1). This analysis extended to lead-specific Q and ST-T abnormalities across all 12 leads (Supplementary Table 5).

In our 681-participant clinical trial, AiTiALVSD predicted 123 individual LVSD phenotypes, distributed into five clusters: Cluster A (16 individuals) exhibited a predominant sinus rhythm, narrow QRS intervals, notable abnormal Q waves in anterior (V2-4) and septal leads, and a significant T wave inversion in anterior and lateral leads. This cluster, with an 81.25% true positive rate, had a history of hypertension, coronary artery disease, and chronic kidney disease, with average AiTiALVSD scores and LVEF of 42.9 and 36.7%, respectively.

Cluster B (13 ECGs), with the least CAD and CKD prevalence, displayed no ST segment deviations, a common QT prolongation, and ST elevation, with the lowest average AiTiALVSD score of 33.2 and LVEF of 41.4%.

Cluster C (32 ECGs) noted for its high QRS duration and T inversion rate in inferior leads, showed the most ST depressions and the highest LVH interpretation by ECG devices, alongside the longest E/E’ and E/A ratios. This cluster had the highest AiTiALVSD score of 55.87 with an LVEF of 35.97%.

Cluster D (49 ECGs), characterized by high CAD prevalence, had the lowest mean heart rate, shortest QTc interval, presence of pacemakers, and similar Q wave abnormalities to Cluster A, with average LVEF and AiTiALVSD scores of 39.3% and 40.0, respectively.

Cluster E (13 individuals) showed the lowest prevalence of past medical conditions, highest heart rate, shortest QRS duration, highest RAD occurrence, and significant AF presence, with average LVEF and AiTiALVSD scores of 38.1% and 37.7, respectively. Supplementary Figures 5 through 9 display representative true positive ECGs for each cluster.

### Lead reversal analysis

Within the 681-case full analysis set, 12 unique ECG lead reversal configurations were examined, with performance outcomes detailed in Table 3. AiTiALVSD consistently demonstrated high diagnostic accuracy, achieving an AUROC of over 0.9 for all but the ‘RA/LL reversal and Counter clockwise reversal without LA’ scenario. Furthermore, the model maintained a sensitivity rate above 80% across these conditions, highlighting its reliable detection capabilities even amidst lead reversals.

**Table 3.** Diagnostic performance of AiTiALVSD; 12 types of lead reversal ECGs.

|  | AUROC<br>(95% CI) | AUPRC<br>(95% CI) | Sensitivity<br>(95% CI) | Specificity<br>(95% CI) | PPV<br>(95% CI) | NPV<br>(95% CI) |
| --- | --- | --- | --- | --- | --- | --- |
| 1. [Nonreversal] or [LL/RL reversal] | 0.971<br>(0.954-0.989) | 0.883<br>(0.838-0.923) | 0.898<br>(0.838-0.958) | 0.940<br>(0.921-0.959) | 0.715<br>(0.636-0.795) | 0.982<br>(0.971-0.993) |
| 2. [LL/LA reversal] or [Counter clockwise reversal w/o RA] | 0.965<br>(0.950-0.980) | 0.834<br>(0.774-0.885) | 0.908<br>(0.851-0.965) | 0.892<br>(0.867-0.917) | 0.586<br>(0.507-0.664) | 0.983<br>(0.972-0.994) |
| 3. [Clockwise w/o RA] or [RL/LA reversal] | 0.966<br>(0.951-0.980) | 0.824<br>(0.760-0.883) | 0.867<br>(0.800-0.935) | 0.925<br>(0.903-0.946) | 0.659<br>(0.577-0.741) | 0.976<br>(0.964-0.989) |
| 4. [RA/LA reversal] or [RA/LA and RL/LL reversal] | 0.940<br>(0.912-0.967) | 0.811<br>(0.751-0.866) | 0.827<br>(0.752-0.901) | 0.938<br>(0.919-0.958) | 0.692<br>(0.609-0.776) | 0.970<br>(0.956-0.984) |
| 5. [Clockwise reversal w/o RL] or [Counter clockwise] | 0.921<br>(0.891-0.952) | 0.724<br>(0.647-0.795) | 0.847<br>(0.776-0.918) | 0.870<br>(0.842-0.897) | 0.522<br>(0.444-0.600) | 0.971<br>(0.957-0.986) |
| 6. [RA>LA, LA>RL, LL>RA, RL>LL] or [Clockwise reversal w/o LL] | 0.949<br>(0.929-0.969) | 0.788<br>(0.726-0.851) | 0.806<br>(0.728-0.884) | 0.931<br>(0.911-0.952) | 0.664<br>(0.579-0.749) | 0.966<br>(0.951-0.981) |
| 7. [RA/LL reversal] or [Counter clockwise reversal w/o LA] | 0.893<br>(0.848-0.937) | 0.741<br>(0.667-0.806) | 0.888<br>(0.825-0.950) | 0.708<br>(0.672-0.745) | 0.339<br>(0.281-0.396) | 0.974<br>(0.959-0.989) |
| 8. [Clockwise reversal w/o RL] or [RA>LL, LA>RA, LL>RL, RL>LA] | 0.910<br>(0.874-0.945) | 0.735<br>(0.667-0.798) | 0.939<br>(0.891-0.986) | 0.611<br>(0.571-0.650) | 0.288<br>(0.239-0.338) | 0.983<br>(0.970-0.997) |
| 9. [RA>LL, LA>RL, LL>LA, RL>RA] or [RA/LL and LA/RL reversal] | 0.903<br>(0.864-0.943) | 0.730<br>(0.656-0.801) | 0.898<br>(0.838-0.958) | 0.698<br>(0.661-0.735) | 0.333<br>(0.276-0.390) | 0.976<br>(0.961-0.991) |
| 10. [RA/RL reversal] or [Clockwise reversal w/o LA] | 0.942<br>(0.916-0.968) | 0.816<br>(0.758-0.868) | 0.918<br>(0.864-0.973) | 0.822<br>(0.791-0.853) | 0.464<br>(0.394-0.534) | 0.984<br>(0.972-0.995) |
| 11. [RA/RL and LA/LL reversal] or [RA>RA, LA>LL, LL>RA, RL>LA reversal] | 0.923<br>(0.892-0.954) | 0.744<br>(0.664-0.812) | 0.929<br>(0.878-0.980) | 0.695<br>(0.657-0.732) | 0.338<br>(0.282-0.395) | 0.983<br>(0.971-0.995) |
| 12. [Clockwise reversal] or [Clockwise reversal w/o LL] | 0.932<br>(0.903-0.962) | 0.797<br>(0.734-0.854) | 0.857<br>(0.788-0.926) | 0.883<br>(0.857-0.909) | 0.553<br>(0.474-0.632) | 0.974<br>(0.960-0.987) |

LA, left arm; RA, right arm; LL, left leg; RL, right leg; w/o, without

### Noise robustness analysis

Table 4 presents AiTiALVSD’s performance under various noise influences, such as power line interference, baseline wander and shift, partial white noise, and time mask, demonstrating resilience by showing statistical consistency with clean ECG data, as indicated by overlapping 95% CI across performance metrics. However, exposure to EMG noise revealed a notable decline in specificity (decreasing by 0.088) and PPV (decreasing by 0.197), signaling a reduced ability to accurately identify negative cases and maintain precision in positive predictions under such noise conditions.

**Table 4.**
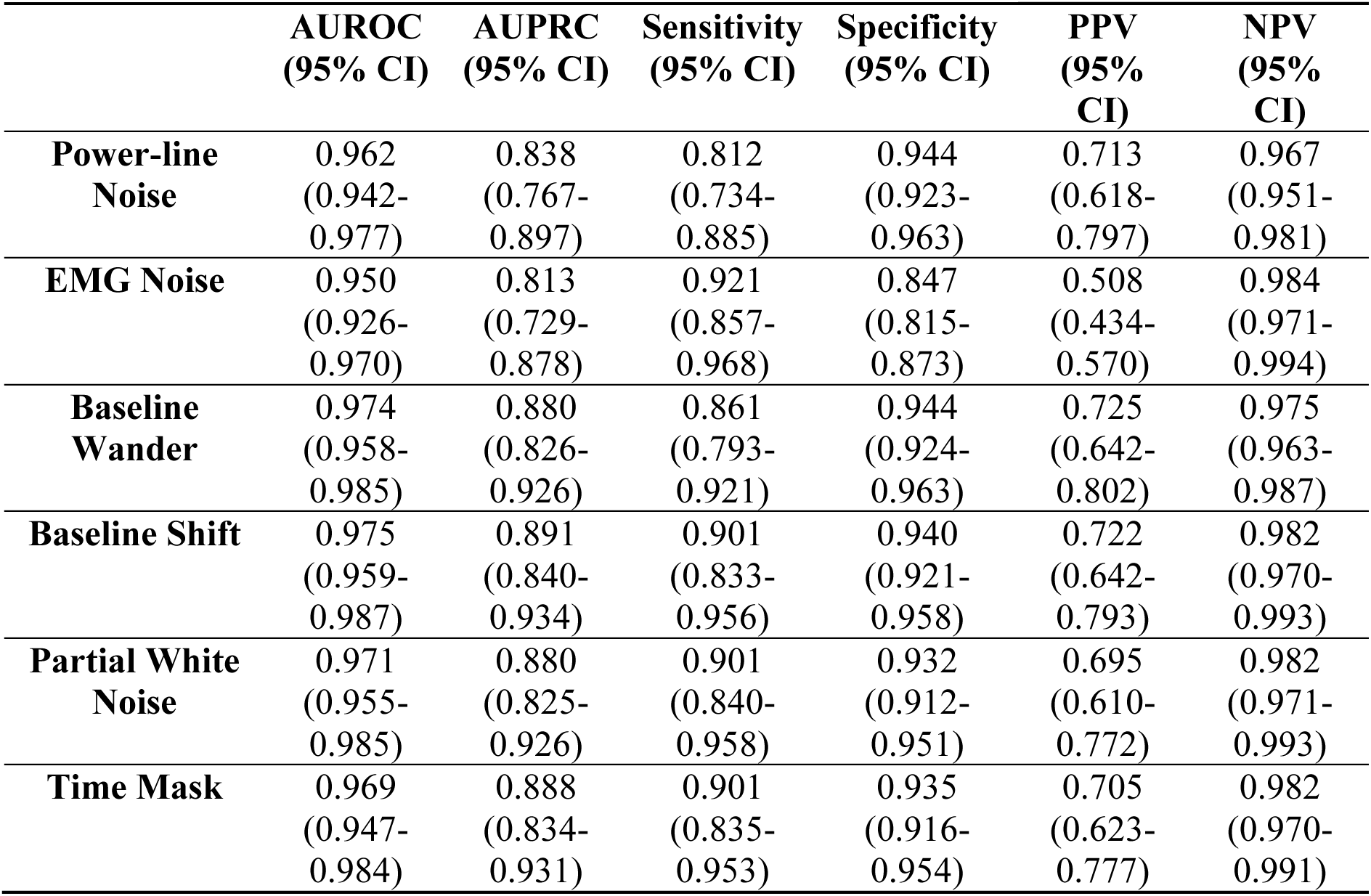
Diagnostic performance of AiTiALVSD; 6 types of ECG noise.

|  | **AUROC**<br>**(95% CI)** | **AUPRC**<br>**(95% CI)** | **Sensitivity**<br>**(95% CI)** | **Specificity**<br>**(95% CI)** | **PPV**<br>**(95%**<br>**CI)** | **NPV**<br>**(95%**<br>**CI)** |
| --- | --- | --- | --- | --- | --- | --- |
| **Power-line**<br>**Noise** | 0.962<br>(0.942-0.977) | 0.838<br>(0.767-0.897) | 0.812<br>(0.734-0.885) | 0.944<br>(0.923-0.963) | 0.713<br>(0.618-0.797) | 0.967<br>(0.951-0.981) |
| **EMG Noise** | 0.950<br>(0.926-0.970) | 0.813<br>(0.729-0.878) | 0.921<br>(0.857-0.968) | 0.847<br>(0.815-0.873) | 0.508<br>(0.434-0.570) | 0.984<br>(0.971-0.994) |
| **Baseline**<br>**Wander** | 0.974<br>(0.958-0.985) | 0.880<br>(0.826-0.926) | 0.861<br>(0.793-0.921) | 0.944<br>(0.924-0.963) | 0.725<br>(0.642-0.802) | 0.975<br>(0.963-0.987) |
| **Baseline Shift** | 0.975<br>(0.959-0.987) | 0.891<br>(0.840-0.934) | 0.901<br>(0.833-0.956) | 0.940<br>(0.921-0.958) | 0.722<br>(0.642-0.793) | 0.982<br>(0.970-0.993) |
| **Partial White**<br>**Noise** | 0.971<br>(0.955-0.985) | 0.880<br>(0.825-0.926) | 0.901<br>(0.840-0.958) | 0.932<br>(0.912-0.951) | 0.695<br>(0.610-0.772) | 0.982<br>(0.971-0.993) |
| **Time Mask** | 0.969<br>(0.947-0.984) | 0.888<br>(0.834-0.931) | 0.901<br>(0.835-0.953) | 0.935<br>(0.916-0.954) | 0.705<br>(0.623-0.777) | 0.982<br>(0.970-0.991) |

## Discussion

The study aimed to investigate the clinical utility of applying transparency and robustness in AI-ECG diagnostics. To the best of our knowledge, this is the first study to apply transparency and robustness principles to AI-ECG diagnostics, marking our initiative as a pioneering effort in this context. The main findings of our research can be summarized as follows: Firstly, The AI-ECG model exhibited substantial diagnostic precision for LVSD, achieving an AUROC of 0.919 (95% CI 0.887-0.951).^10^ Secondly, through TCAV and clustering analyses, the model’s concordance with established medical insights was established, affirming AI-ECG’s capability to accurately reflect complex cardiac conditions. Thirdly, robustness evaluations, encompassing lead reversal and noise tests, demonstrated the model’s steadfast performance in diverse clinical settings, underlining its adaptability and dependability for practical use. This comprehensive approach advances our understanding of AI-ECG’s potential in cardiac care and sets a new benchmark for integrating transparency and robustness in medical diagnostics.

### Model performance

The AI-ECG model demonstrated robust LVSD detection, evidenced by an impressive AUROC of 0.919 for binary outputs and 0.971 for continuous outputs. This validation approach, favoring clinical applicability, introduces binary cutoffs that resonate with healthcare professionals’ operational needs. The model effectively identified LV dysfunction below an LVEF of 40%, with higher AiTiALVSD scores indicating increased LVSD likelihood. Scores exceeding 53.5 were uniquely indicative of LVSD, while scores around the 9.7 cutoff were associated with mild LVEF reduction, notably with over half of the false positives showing an LVEF in the 40-50% range. This nuanced analysis highlights the model’s diagnostic acumen and clinical utility.

Subsequent subgroup analysis on 96 patients with available NT-proBNP data showcased AiTiALVSD’s superior diagnostic accuracy, with a sensitivity of 92.6% surpassing NT-proBNP’s 88.9%. This comparison not only underscores AiTiALVSD’s refined precision but also supports its role as an adjunct tool in early LVSD detection. These insights solidify the merit of AI-enhanced diagnostics over traditional markers like NT-proBNP, reinforcing AI-ECG’s value in cardiovascular screening within both research and practical healthcare domains.^16–18^

### Model transparency

Traditional AI-ECG analysis, predominantly using SHapley Additive exPlanations (SHAP) for basic feature analysis or visual tools like GradCAM for localized insights, often struggles with the depth of explainability required for clinical application, particularly with deep learning models handling extensive datasets.^19,20^ Such methods offer limited clinical insight, underscoring a need for a more holistic interpretative approach.

Addressing this gap, our study integrates advanced methodologies such as TCAV and clustering analysis.^21,22^ This strategy not only aligns the AI model’s functionality with established medical knowledge but also ensures global interpretability crucial for practical medical use. Through TCAV, the model’s reliance on critical ECG concepts for LVSD prediction—like QRS prolongation and abnormal ECG findings—is elucidated, significantly improving transparency and overcoming the conventional black-box critique.

Further, our clustering analysis distinguished five unique LVSD-positive ECG clusters, demonstrating the syndrome’s variability and affirming the model’s capacity to grasp LVSD’s complexity in a manner consistent with medical understanding. This level of detail, showcased through transparent, visual representation, validates AiTiALVSD’s ability to navigate LVSD’s nuances, reinforcing the model’s clinical relevance and trustworthiness.^16^ Transparency in the model’s development process, as detailed in Supplementary Text 2, further establishes a foundation for trust in AI-ECG diagnostics.

### Model Robustness

Transitioning AI-ECG models like AiTiALVSD from research frameworks to clinical environments necessitates a thorough evaluation of their resilience against common practical challenges, including inaccuracies in lead placement and environmental noise. Our extensive robustness assessment of AiTiALVSD, through lead reversal simulations in a 12-lead ECG configuration and exposure to diverse noise conditions, showcased the model’s remarkable resilience. AiTiALVSD achieved an AUROC of 0.893 to 0.971 and sensitivity rates from 80.6% to 92.9% across lead reversal conditions, and an AUROC of 0.950 to 0.975 with sensitivities ranging from 81.2% to 92.1% amidst various noise types. These results underscore AiTiALVSD’s steady and reliable diagnostic capabilities in adverse settings. Such comprehensive evaluation not only underscores AiTiALVSD’s clinical deployment readiness but also establishes a new robustness benchmark for AI-ECG models, paving the way for future advancements in the discipline.

### Limitations

Our study’s insights come with inherent constraints that necessitate cautious interpretation. The retrospective design and single-center focus within South Korea may not capture the global population diversity, suggesting careful extrapolation to other demographics. Future studies should seek to include broader, multi-ethnic samples to ensure wider applicability and enhance the findings’ generalizability. Moreover, while the model displayed 80-90% sensitivity at a 9.7 binary cutoff, this threshold’s universality across diverse cohorts remains uncertain. The model’s adaptability, akin to traditional biomarkers, underscores the necessity for ongoing validation in varied clinical settings to affirm its reliability and broader clinical utility.

Additionally, the retrospective nature introduces potential selection and information biases, highlighting the importance of future prospective studies and randomized trials to underpin the model’s efficacy robustly. A notable constraint is our limited sample size, which curtailed our capacity to fully articulate the electrocardiographic and clinical nuances within identified clusters, limiting the depth of prognostic or long-term outcome analyses. Subsequent research should aim for larger cohorts to facilitate a more granular exploration of these clusters, including longitudinal studies to discern specific prognostic indicators and outcomes, thus amplifying our findings’ clinical relevance.

### Conclusion

In conclusion, this study confirms the effectiveness of our AI-ECG model in detecting LVSD, emphasizing its accuracy, transparency, and robustness. It suggests the importance of using TCAV for comprehensive interpretation and conducting noise and lead reversal resilience tests. This research sets a new benchmark in AI-ECG diagnostics, highlighting the critical role of methodological rigor in enhancing early LVSD detection.

## Methods

### Study design

This retrospective, single-center cohort study analyzed data from a pivotal clinical study on AI/ML-enabled Software as a Medical Device (SaMD) for ECG analysis, named AiTiALVSD. The study’s objective was to evaluate AiTiALVSD’s diagnostic performance, transparency, and robustness by juxtaposing ECG analysis outcomes with echocardiographic data, the reference standard, in patients 18 years or older suspected of LVSD at Mediplex Sejong Hospital in the Republic of Korea, spanning March 2017 to August 2021. Figure 1 shows study flow for evaluating ECG SaMD.

Inclusion criteria required a 12-lead ECG taken within 24 hours prior to echocardiography, a 10-second measurement duration, and a sampling rate of 500 Hz using PAGEWRITER TC30 or TC70 (Philips, USA). If multiple ECGs were available, the one closest in time to the echocardiography was chosen. Exclusion criteria included missing ECG data, gaps in recording, or lack of echocardiography to confirm left ventricular ejection fraction (LVEF). This study received approval for the investigational device exemption (IDE) trial from the institutional review boards of Mediplex Sejong Hospital (IRB File No. ISH-2022-09-005-003), with informed consent waived due to its retrospective study design.

### Variables and reference standards

Patient demographic and clinical profiles, including age, sex, baseline characteristics, and lab results, were retrieved from electronic records, with LVEF assessed via the biplane Simpson method. LVSD was identified with an EF ≤ 40%, contrasting a non-LVSD definition of EF > 40%. Reference standard data was anonymized to maintain confidentiality. An independent evaluator, blind to LVSD status, inputted the 12-lead ECG data into AiTiALVSD, documenting the algorithm-determined LVSD scores and risk categories.

### AI-ECG model description

AiTiALVSD, an AI/ML-based SaMD, exclusively analyzes 12-lead ECG raw data to aid in diagnosing LVSD, determining if the LVEF is 40% or lower. Upon processing ECG data, it outputs a 0-100 score reflecting LVSD likelihood. Scores ≥9.7, set for 90% sensitivity from prior research, indicate high LVSD probability, while scores below this mark suggest low risk.^7^

Developed around a one-dimensional residual neural network (ResNet) using the PyTorch framework and Python, AiTiALVSD features a composition of a stem block, four feature blocks, and a fully connected layer for LVSD pattern recognition.^23,24^ Each feature block, a series of three residual blocks, sequentially processes and extracts ECG characteristics for analysis. The model inputs digital 12-lead ECGs, captured at a 500 Hz sampling rate over 10 seconds, and classifies outcomes as binary LVSD indications. Further model development details and data distribution specifics are available in Supplementary Texts 1 and 2.

### Experiments for Transparency

In the clinical domain, significance of explainable AI has escalated, emphasizing two critical aspects: clinical plausibility and exploitation.^21^ Clinical plausibility confirms that AI-generated outcomes are reliable and relevant in clinical environments. In contrast, exploitation refers to the application of these findings to corroborate existing medical insights. Whereas previous research predominantly concentrated on local interpretative methods, which clarify the rationale behind AI predictions for singular data points by spotlighting key features, this approach inadequately addresses the explanation of specific characteristics, Conversely, global interpretative strategies strive to demystify the overarching rationale of AI models across diverse inputs, providing an integral perspective of their predictive logic.^17,18^

#### 1. Testing with Concept Activation Vectors (TCAV)

The implementation of TCAV allows for the assessment of AiTiALVSD’s consistency with medically recognizable concepts. TCAV scores, derived from this evaluation, quantify the influence of specific concepts on AiTiALVSD’s feature blocks during prediction. These scores, ranging from 0 to 1, signify the degree of positive impact a concept has on predicting LVSD, with scores near 1 indicating significant influence. Utilizing the Physionet Challenge 2021 dataset^25^, we constructed nine “LVSD concept” datasets from 26 different ECG interpretation labels: atrial fibrillation and atrial flutter (AF), left bundle branch block (LBBB), right bundle branch block (RBBB), left axis deviation (LAD), right axis deviation (RAD), prolonged QT, conduction disorder, abnormal Q wave and abnormal T wave. The AF concept included both atrial fibrillation and atrial flutter. The abnormal T wave concept included both abnormal T wave and T wave inversion labels. These “LVSD concepts” represent ECG indicators recognized as heart failure with reduced ejection fraction risk factors, supported by recent studies.^26,27^ TCAV scores are presented with a 95% confidence interval (CI) for precision, detailed further in Supplementary Text 3. For analysis, we utilized the captum python package (version 0.6.0)^28^, with TCAV research code for ECG available at: https://github.com/medicalai-research/TCAV_ECG.

#### 2. Cluster analysis

Our goal was to phenotype LVSD by clustering AiTiALVSD’s hidden features, especially those within the block demonstrating the highest TCAV score. We posited that these prominent features would most accurately reflect ECG characteristics in alignment with established medical knowledge. Through a comparative analysis of demographic, electrocardiographic, and echocardiographic features across clusters, we verified AiTiALVSD’s capability to discern key LVSD traits. Employing positively predicted ECGs from a previous development dataset, we trained a K-means clustering algorithm, subsequently applying the model to our dataset. The optimal cluster count was ascertained using the elbow algorithm.^29^ Detailed methodologies are elaborated in Supplementary Text 3.

##### Tests for robustness

Acknowledging the quality variance between data from controlled experiments and real-world clinical scenarios, we initiated robustness assessments to preemptively identify and mitigate potential model performance discrepancies due to data quality changes.

#### 1. Lead reversal test

To assess our model’s resilience against electrode misplacement, we generated potential 12-lead ECG morphologies for each patient, resulting in an evaluation of the model across 12 distinct types of lead reversal scenarios from a pool of 681 ECGs.^30^ Despite the 24 conceivable lead reversal permutations, only 12 unique morphologies were identified due to identical presentations in certain pairs, e.g., [Nonreversal] or [LL/RL reversal], highlighting the model’s diagnostic accuracy in identifying LVSD under varied electrode configurations.

#### 2. Noise robustness test

Our noise resilience test incorporated six types of artificial noise: power line, electromyography (EMG), baseline wander and shift, partial white noise, and time mask noise, reflecting common distortions in real ECG data. By applying these noise profiles to our dataset, we observed AiTiALVSD’s performance shifts, identifying critical noise factors impacting accuracy. This comprehensive approach enables the determination of AiTiALVSD’s noise robustness, as detailed in Supplementary Figure 1 and validated across the modified datasets.

### Statistical analysis

Data were analyzed using statistical tests, such as Student’s t test, Mann–Whitney U test, chi-square test, and Fisher’s exact test, as appropriate. Correlation analysis was performed using Pearson correlation methods. Model performance was assessed by calculating the AUROC. We determined the AiTiALVSD threshold point that gave a sensitivity ≥90% (AiTiALVSD score 9.7) during model development. We report this threshold’s sensitivity, specificity, negative predictive value (NPV), and positive predictive value (PPV), along with 95% CI. We confirmed the AUROC with both continuous predictors and binary predictors. AUPRC was also estimated to the model performance, using only a continuous output variable. For the lead reversal and noise robustness analyses, AUROC and AUPRC were estimated based on the continuous output variable. Subgroup analyses were performed by demographics, medical history, several ECG patterns, and NT-proBNP. To adhere to the conditions set by Mediplex Sejong Hospital, patients with NT-pro BNP test results equal to or greater than 158 pg/mL (ARCHITECT i2000 SR, Abbott) were classified as abnormal, while those with results below 158 pg/mL were considered normal.

In this study, we utilized AiTiALVSD version 1.00.00 (Medical AI Co., Ltd., Seoul, Republic of Korea) aimed at obtaining approval from the Korea Food and Drug Administration (KFDA) for a SaMD. The study was designed to meet the pivotal standards required for SaMD certification, with the performance of AiTiALVSD anticipated at an AUROC of 0.85. The criterion for clinical validity evaluation was set at an AUROC of 0.75, with the significance level (α) determined at 0.05 and the statistical power (1-β) targeted at 95%, ensuring compliance with the stringent regulatory requirements for medical device approval in South Korea. Considering a dropout rate of 5%, the total sample size was 688 participants (103 LVSD and 585 Non-LVSD). We elaborated thorough backgrounds for the sample size calculation in the supplementary Text 4. Sample size calculation was performed using PASS 15 (NCSS, LLC. Kaysville, USA) software. All analyses were conducted using R (version 4.2.3) and Python (version 3.8).

## Data Availability

The data referred to in this manuscript are proprietary to the hospital and, as such, cannot be shared publicly in accordance with institutional policies and privacy regulations.

## Abbreviations

AF: atrial fibrillation
AI: artificial intelligence
CI: confidence interval
ECG: electrocardiogram
EF: ejection fraction
EMG: electromyography
HF: heart failure
IDE: investigational device exemption
LAD: left axis deviation
LBBB: left bundle branch block
LVSD: left ventricular systolic dysfunction
NPV: negative predictive value
PPV: positive predictive value
RAD: right axis deviation
RBBB: right bundle branch block
ResNet: residual neural network
SaMD: software as a medical device
TCAV: testing with concept activation vectors

## Acknowledgments

None

## Central illustration

To evaluate the transparency and robustness of the AI-ECG model, we applied the following four methods: Testing with Concept Activation Vectors (TCAV), clustering analysis, ECG noise tests, and lead reversal tests.

