## Supplementary for "Transparent and robust Artificial intelligence-driven Electrocardiogram model for Left Ventricular Systolic Dysfunction"

**Supplemental Content**

| ***e*Methods** |  |
| --- | --- |
| **Supplemental Text 1.** | AiTiALVSD algorithm development |
| **Supplemental Text 3.** | Interpretation methods for model transparency |
| ***e*Results** |  |
| **Supplemental Figure 1.** | An example of ECG noise type. |
| Supplemental Figure 2. | AiTiALVSD score distribution. |
| **Supplemental Figure 3.** | Pearson’s correlation analysis using the AiTiALVSD score and LVEF |
| **Supplemental Figure 4.** | Forest plots of subgroup analyses |
| **Supplemental Figure 5-9.** | An example ECG of cluster A, B, C, D and E. |
| **Supplemental Table 1.** | Tertiary outcomes |
| **Supplemental Table 2.** | AiTiALVSD performance by subgroup |
| **Supplemental Table 3.** | The performance of AiTiALVSD using a continuous output variable |
| **Supplemental Table 4.** | Performance comparison between NT-proBNP and AiTiALVSD among 96 patients |
| **Supplemental Table 5.** | Phenotyping of AiTiALVSD-positive LVSD cases |
| **Supplemental Text 2.** | Model Card of AiTiALVSD for model development |

**Supplemental Figures**

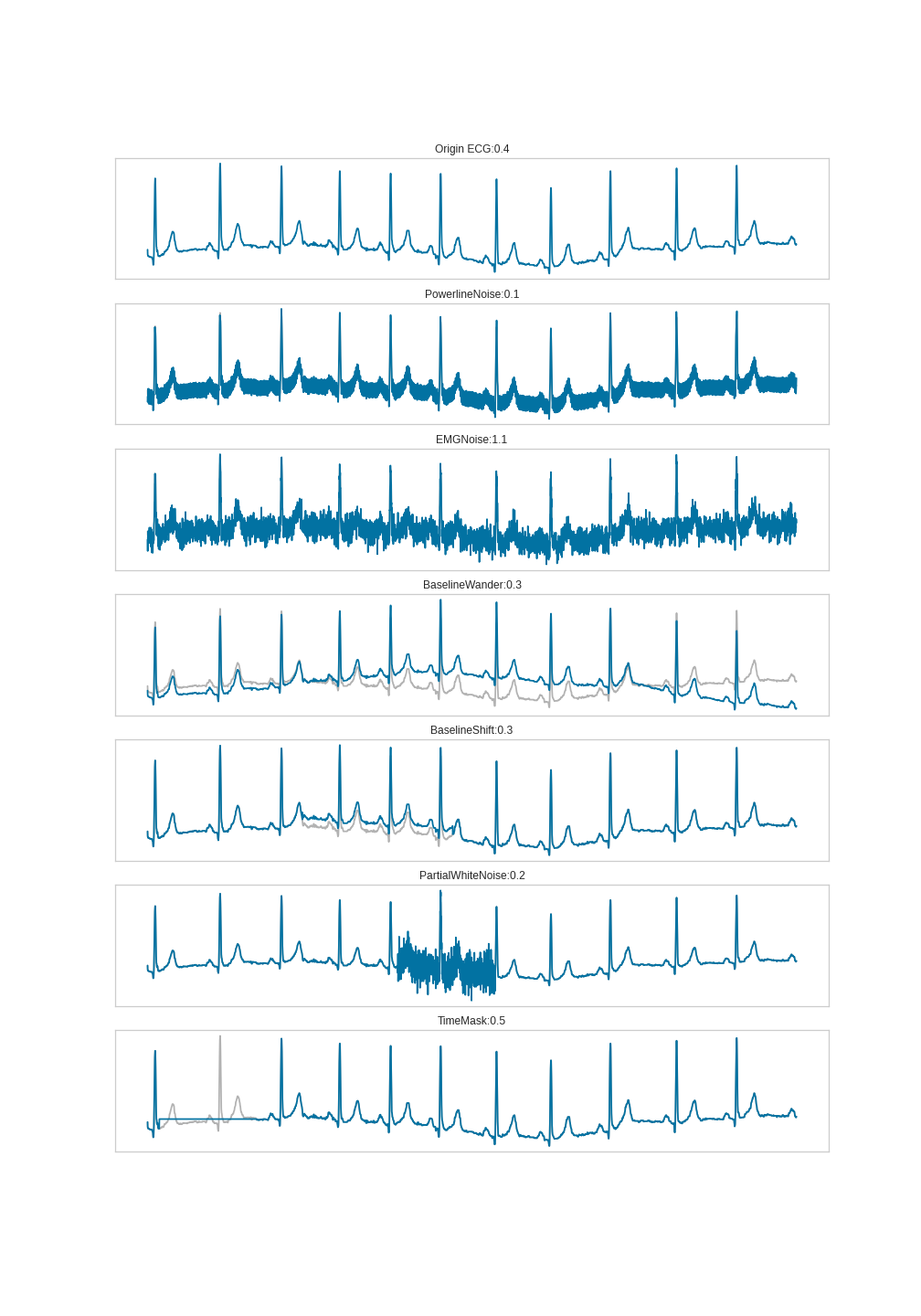

**Supplemental Figure 1.** An example of ECG noise type. Each score is the AiTiALVSD score when augmented ECG (blue) with each type of noise is predicted. Gray-colored ECG is an original ECG.

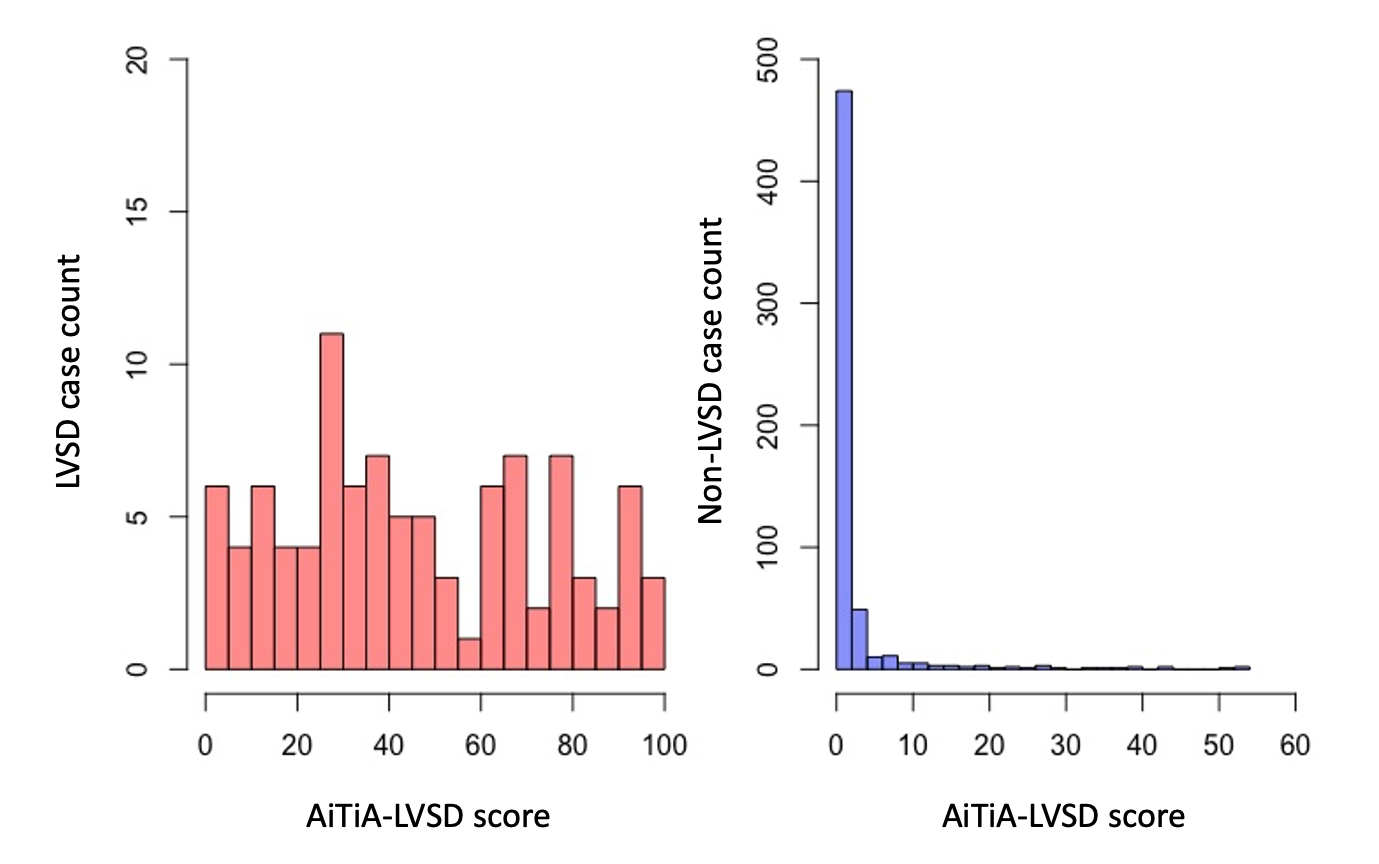

Supplemental Figure 2. AiTiALVSD score distribution. When the score distribution in the LVSD and Non-LVSD groups is expressed as a histogram, the two groups are clearly distributed with scores above and below 9.7

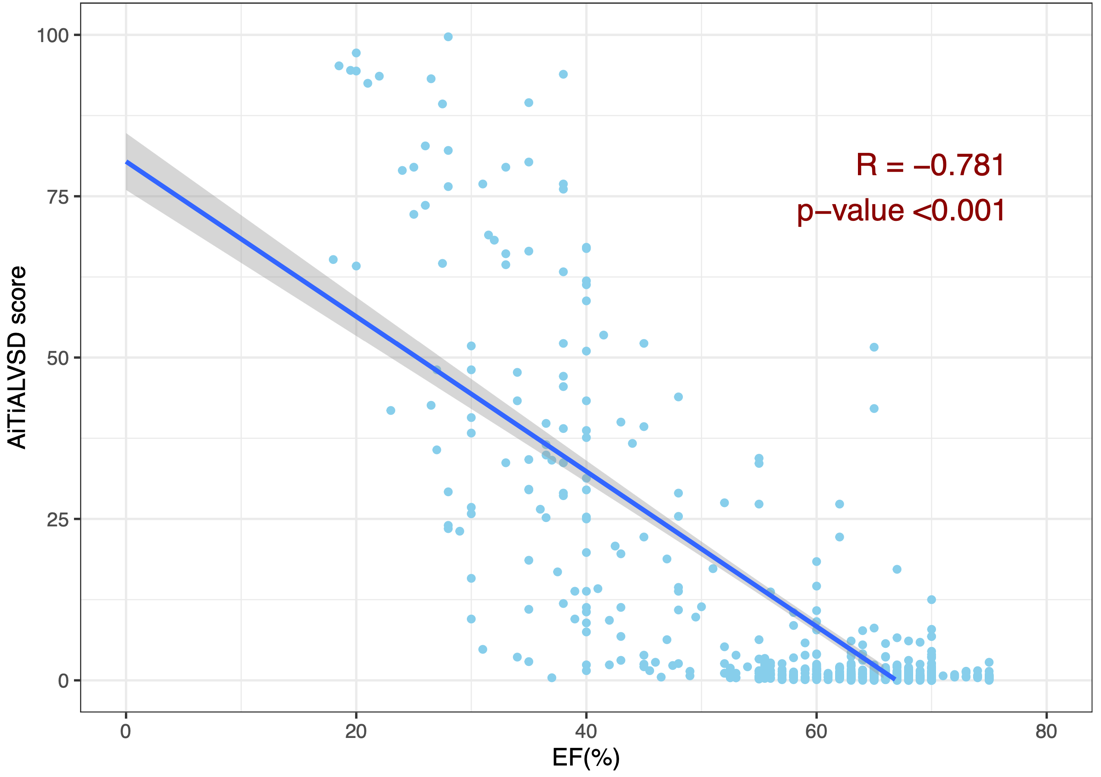

**Supplemental Figure 3.** Pearson’s correlation analysis using the AiTiALVSD score and LVEF

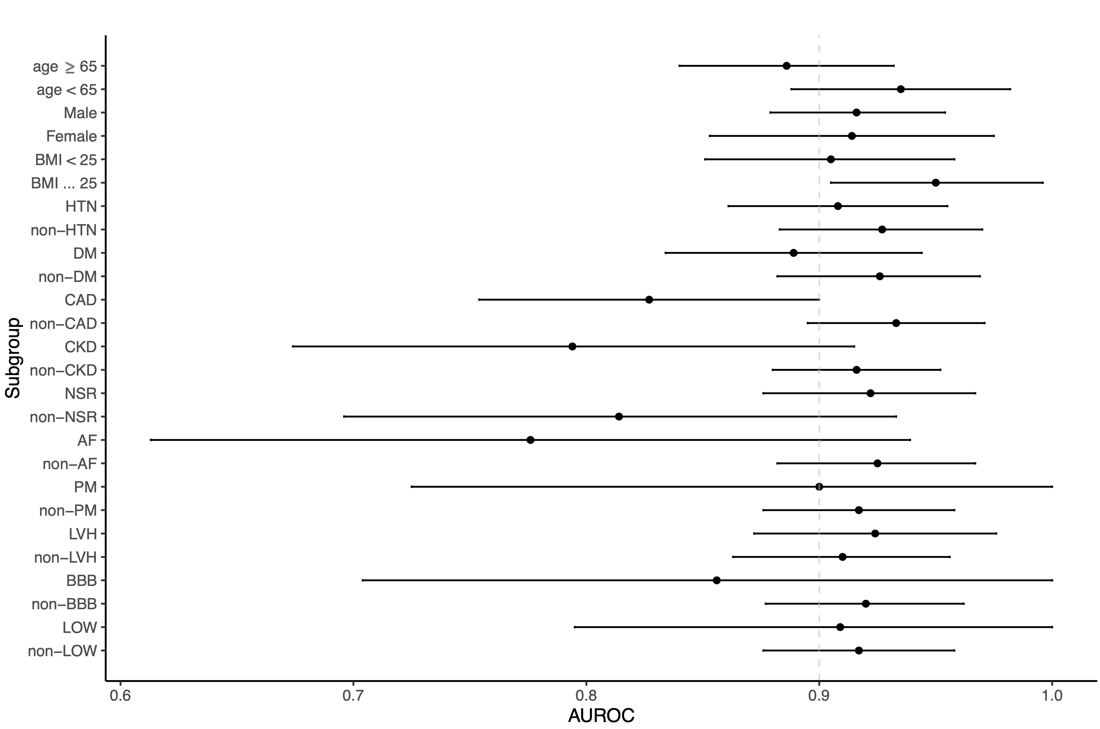

**Supplemental Figure 4. Forest plots of subgroup analyses**

The AUROC and 95% confidence interval of AiTiALVSD in subgroups according to baseline characteristics and ECG interpretation are displayed in a forest plot; it shows overall performance at 0.8 to 0.9. BMI, body mass index; HTN, hypertension; DM, diabetes mellitus; CAD, coronary artery disease; CKD, chronic kidney disease; SR, sinus rhythm; AF, atrial fibrillation; PM, pacemaker; LVH, left ventricular hypertrophy; BBB, right and left bundle branch block; LOW, low voltage

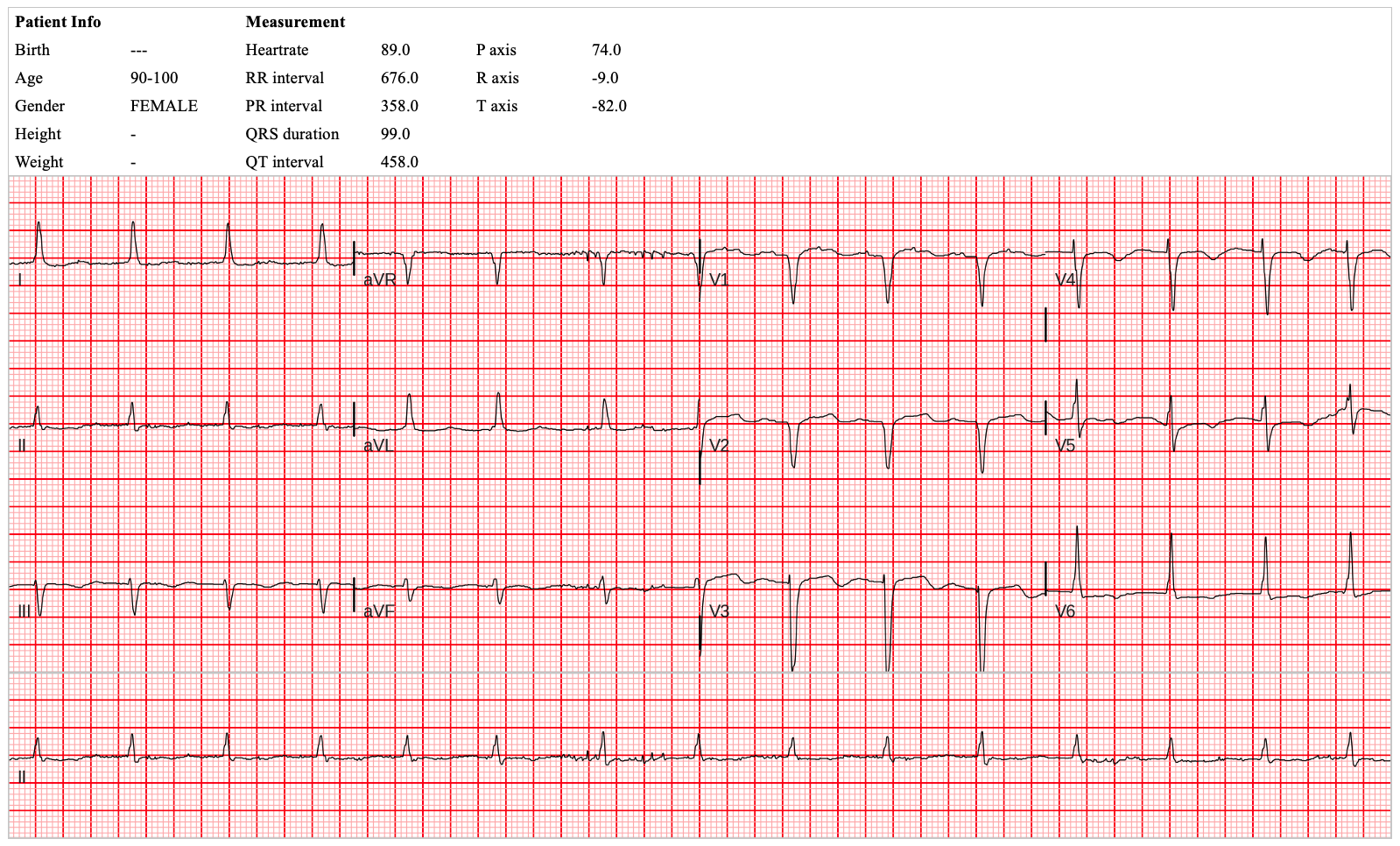

**Supplemental Figure 5.** An example ECG of cluster A

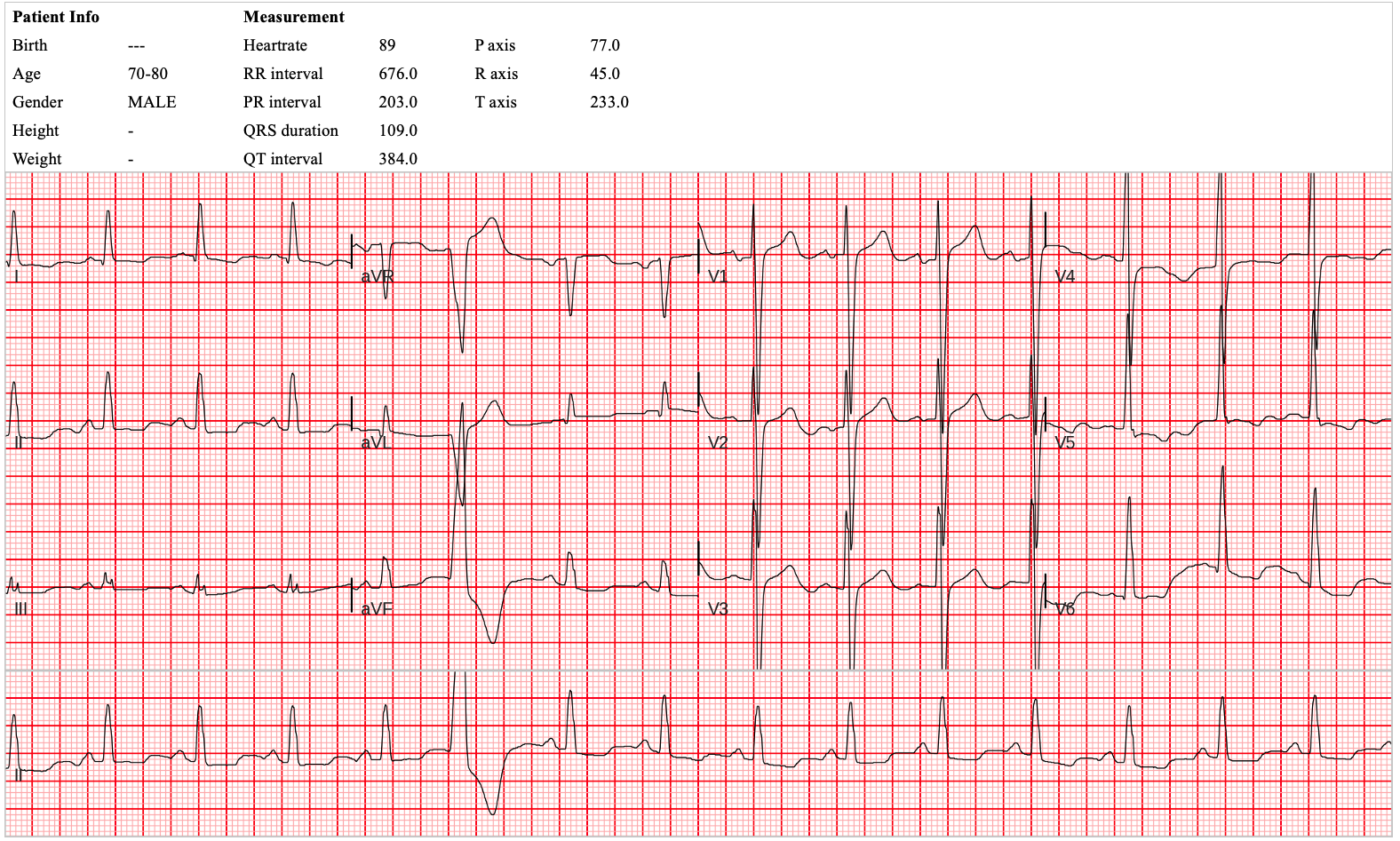

**Supplemental Figure 6.** An example ECG of cluster B

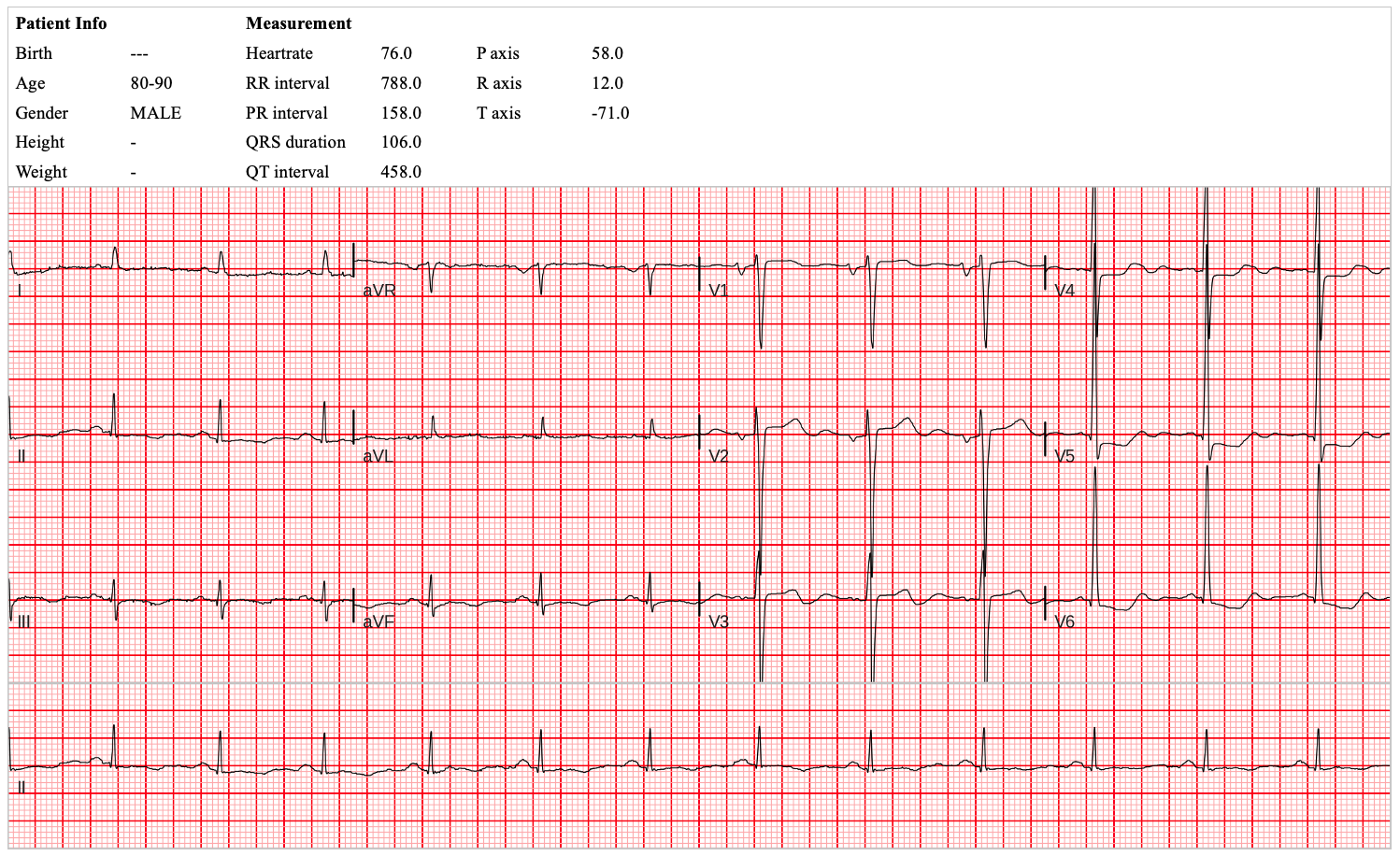

**Supplemental Figure 7.** An example ECG of cluster C

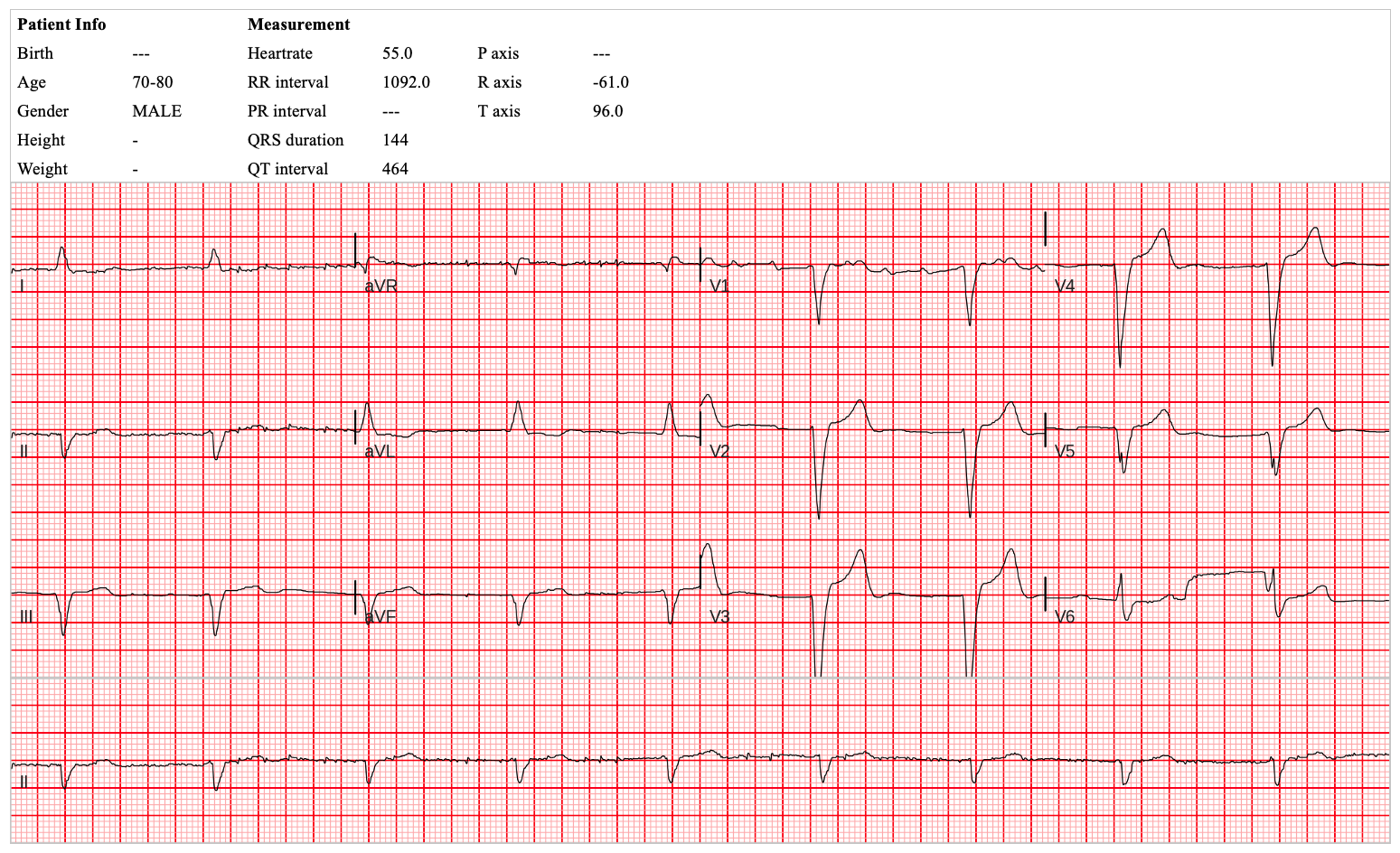

**Supplemental Figure 8.** An example ECG of cluster D

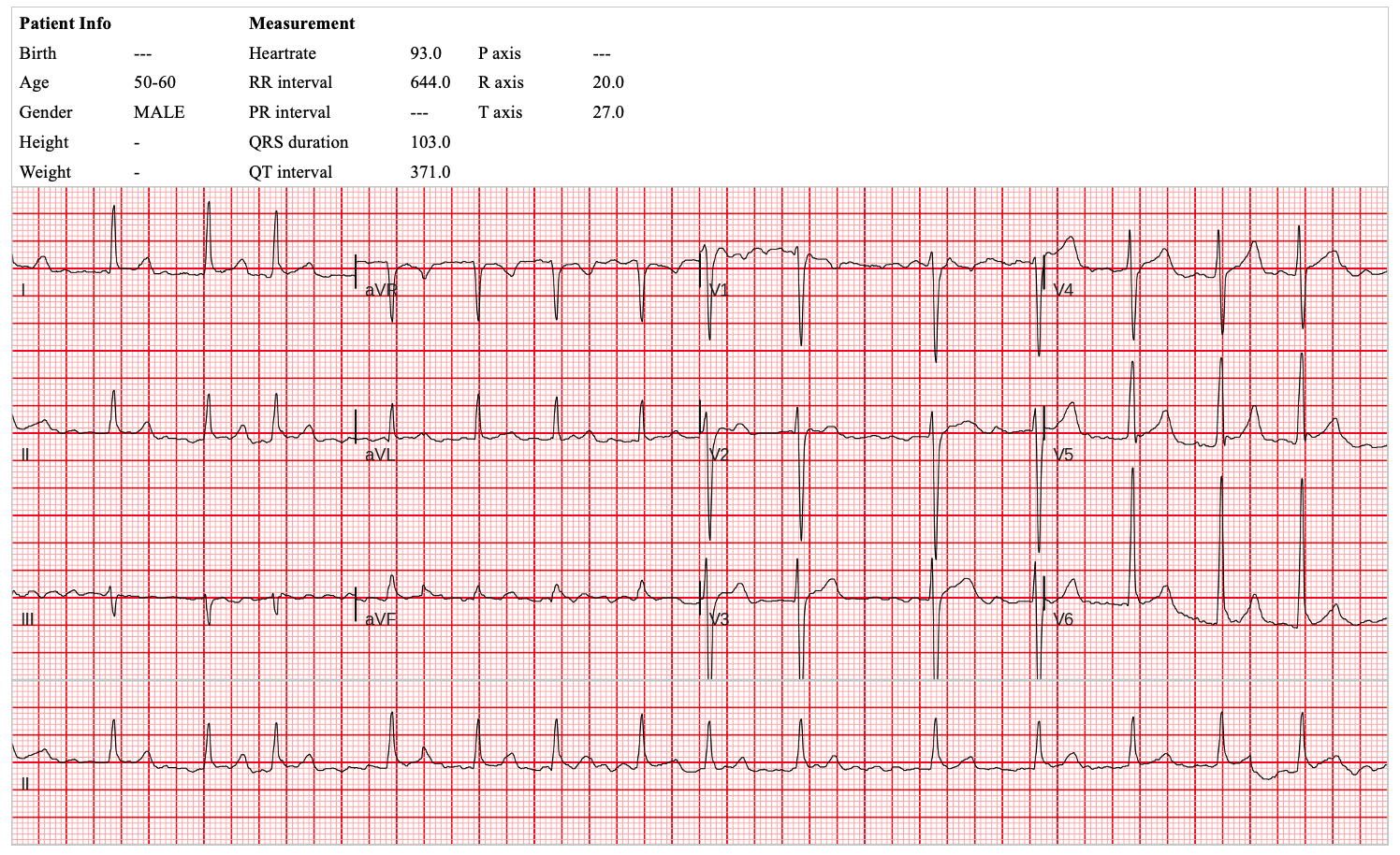

**Supplemental Figure 9.** An example ECG of cluster E

**Supplemental Tables**

**Supplemental Table 1.** Tertiary outcomes

|  | AUROC  (95% CI) | Sensitivity  (95% CI) | Specificity  (95% CI) | PPV  (95% CI) | NPV  (95% CI) |
| --- | --- | --- | --- | --- | --- |
| AiTiALVSD   (LVEF  ≤ 35%) | 0.906 | 0.927 | 0.885 | 0.415 | 0.993 |
|  | (0.869-0.943) | (0.859-0.996) | (0.860-0.910) | (0.328-0.502) | (0.986-1.0) |
| AiTiALVSD   (LVEF ≤ 50%) | 0.891 | 0.811 | 0.971 | 0.870 | 0.955 |
|  | (0.857-0.925) | (0.744-0.877) | (0.957-0.985) | (0.810-0.929) | (0.938-0.972) |

LVEF, left ventricular ejection fraction; AUROC, area under the receiver operating characteristic curve; PPV, positive predictive value; NPV, negative predictive value; CI, confidence interval

**Supplemental Table 2.** AiTiALVSD performance by subgroup

|  | AUROC (95% CI) | AUPRC (95% CI) | Sensitivity (95% CI) | Specificity (95% CI) | PPV (95% CI) | NPV (95% CI) |
| --- | --- | --- | --- | --- | --- | --- |
| Age |  |  |  |  |  |  |
| age ≥ 65  (n=263) | 0.886 | 0.591 | 0.906 | 0.867 | 0.632 | 0.973 |
|  | (0.840-0.932) | (0.497-0.683) | (0.827-0.984) | (0.821-0.913) | (0.523-0.740) | (0.950-0.996) |
| age < 65  (n=417) | 0.935 | 0.768 | 0.889 | 0.981 | 0.851 | 0.986 |
|  | (0.888-0.982) | (0.658-0.868) | (0.797-0.981) | (0.967-0.995) | (0.749-0.953) | (0.975-0.998) |
| Sex |  |  |  |  |  |  |
| Male  (n=350) | 0.916 | 0.686 | 0.91 | 0.922 | 0.735 | 0.978 |
|  | (0.879-0.954) | (0.605-0.764) | (0.842-0.979) | (0.891-0.953) | (0.640-0.830) | (0.960-0.995) |
| Female  (n=331) | 0.914 | 0.600 | 0.871 | 0.957 | 0.675 | 0.986 |
|  | (0.853-0.975) | (0.460-0.736) | (0.753-0.989) | (0.934-0.980) | (0.530-0.820) | (0.973-1.0) |
| BMI |  |  |  |  |  |  |
| BMI < 25 (n=396) | 0.905 | 0.679 | 0.864 | 0.945 | 0.76 | 0.972 |
|  | (0.851-0.958) | (0.588-0.759) | (0.781-0.946) | (0.921-0.970) | (0.663-0.857) | (0.954-0.99) |
| BMI ≥ 25 (n=282) | 0.950 | 0.870 | 0.969 | 0.932 | 0.646 | 1.00 |
|  | (0.905-0.996) | (0.780-0.940) | (0.908-1.00) | (0.901-0.963) | (0.511-0.781) | (0.987-1.00) |
| HTN |  |  |  |  |  |  |
| HTN  (n=288) | 0.908 | 0.639 | 0.896 | 0.921 | 0.694 | 0.978 |
|  | (0.861-0.955) | (0.541-0.742) | (0.809-0.982) | (0.887-0.955) | (0.579-0.808) | (0.959-0.997) |
| non-HTN  (n=393) | 0.927 | 0.677 | 0.9 | 0.953 | 0.738 | 0.985 |
|  | (0.883-0.970) | (0.570-0.777) | (0.817-0.983) | (0.931-0.976) | (0.627-0.848) | (0.972-0.998) |
| DM |  |  |  |  |  |  |
| DM  (n=141) | 0.889 | 0.749 | 0.896 | 0.88 | 0.796 | 0.942 |
|  | (0.834-0.944) | (0.646-0.841) | (0.809-0.982) | (0.814-0.947) | (0.689-0.904) | (0.892-0.991) |
| non-DM  (n=540) | 0.926 | 0.596 | 0.9 | 0.951 | 0.652 | 0.989 |
|  | (0.882-0.969) | (0.501-0.690) | (0.817-0.983) | (0.932-0.970) | (0.540-0.765) | (0.980-0.999) |
| CAD |  |  |  |  |  |  |
| CAD  (n=104) | 0.827 | 0.631 | 0.889 | 0.765 | 0.667 | 0.929 |
|  | (0.754-0.90) | (0.514-0.742) | (0.786-0.992) | (0.664-0.866) | (0.533-0.80) | (0.861-0.996) |
| non-CAD  (n=577) | 0.933 | 0.685 | 0.903 | 0.963 | 0.747 | 0.988 |
|  | (0.895-0.971) | (0.583-0.771) | (0.830-0.977) | (0.947-0.979) | (0.648-0.845) | (0.979-0.998) |
| CKD |  |  |  |  |  |  |
| CKD  (n=31) | 0.794 | 0.667 | 1.00 | 0.588 | 0.667 | 1.00 |
|  | (0.674-0.915) | (0.478-0.826) | (1.00-1.00) | (0.354-0.822) | (0.465-0.868) | (1.00-1.00) |
| non-CKD  (n=650) | 0.916 | 0.655 | 0.881 | 0.951 | 0.725 | 0.982 |
|  | (0.880-0.952) | (0.577-0.733) | (0.812-0.950) | (0.933-0.968) | (0.639-0.812) | (0.971-0.993) |
| SR |  |  |  |  |  |  |
| NSR  (n=613) | 0.922 | 0.659 | 0.886 | 0.958 | 0.729 | 0.985 |
|  | (0.876-0.967) | (0.574-0.756) | (0.811-0.960) | (0.941-0.975) | (0.635-0.824) | (0.974-0.995) |
| non-NSR (n=68) | 0.814 | 0.665 | 0.929 | 0.7 | 0.684 | 0.933 |
|  | (0.696-0.933) | (0.542-0.794) | (0.833-1.00) | (0.558-0.842) | (0.536-0.832) | (0.844-1.00) |
| AF |  |  |  |  |  |  |
| AF  (n=43) | 0.776 | 0.670 | 0.9 | 0.652 | 0.692 | 0.882 |
|  | (0.613-0.939) | (0.526-0.818) | (0.769-1.00) | (0.458-0.847) | (0.515-0.870) | (0.729-1.00) |
| non-AF  (n=638) | 0.925 | 0.660 | 0.897 | 0.952 | 0.722 | 0.985 |
|  | (0.882-0.967) | (0.580-0.739) | (0.830-0.965) | (0.934-0.970) | (0.632-0.811) | (0.975-0.995) |
| PM |  |  |  |  |  |  |
| PM (n=10) | 0.9 | 0.833 | 1.00 | 0.8 | 0.833 | 1.00 |
|  | (0.725-1.00) | (0.500-1.00) | (1.00-1.00) | (0.449-1.00) | (0.535-1.00) | (1.00-1.00) |
| non-PM (n=671) | 0.917 | 0.648 | 0.892 | 0.941 | 0.709 | 0.982 |
|  | (0.876-0.958) | (0.571-0.723) | (0.830-0.955) | (0.922-0.960) | (0.627-0.792) | (0.971-0.993) |
| LVH |  |  |  |  |  |  |
| LVH (n=66) | 0.924 | 0.741 | 1.00 | 0.848 | 0.741 | 1.00 |
|  | (0.872-0.976) | (0.600-0.875) | (1.00-1.00) | (0.744-0.952) | (0.575-0.906) | (1.00-1.00) |
| non-LVH (n=615) | 0.91 | 0.634 | 0.872 | 0.948 | 0.708 | 0.981 |
|  | (0.863-0.956) | (0.546-0.712) | (0.798-0.946) | (0.929-0.967) | (0.617-0.799) | (0.969-0.993) |
| BBB |  |  |  |  |  |  |
| BBB  (n=37) | 0.856 | 0.698 | 0.929 | 0.783 | 0.722 | 0.947 |
|  | (0.704-1.00) | (0.503-0.867) | (0.794-1.00) | (0.614-0.951) | (0.515-0.929) | (0.847-1.00) |
| non-BBB (n=644) | 0.92 | 0.652 | 0.893 | 0.946 | 0.714 | 0.983 |
|  | (0.877-0.962) | (0.579-0.726) | (0.827-0.959) | (0.928-0.965) | (0.628-0.801) | (0.972-0.994) |
| LOW |  |  |  |  |  |  |
| LOW (n=16) | 0.909 | 0.714 | 1.00 | 0.818 | 0.714 | 1.00 |
|  | (0.795-1.00) | (0.400-1.00) | (1.00-1.00) | (0.590-1.00) | (0.380-1.00) | (1.00-1.00) |
| non-LOW (n=665) | 0.917 | 0.654 | 0.892 | 0.942 | 0.716 | 0.982 |
|  | (0.876-0.958) | (0.580-0.723) | (0.830-0.955) | (0.923-0.961) | (0.633-0.798) | (0.971-0.993) |

**Supplemental Table 3.** The performance of AiTiALVSD using a continuous output variable

|  | AUROC  (95% CI) | AUPRC  (95% CI) |
| --- | --- | --- |
| LVEF ≤ 40% | 0.971 | 0.883 |
|  | (0.956-0.985) | (0.839-0.925) |
| LVEF ≤ 35% | 0.966 | 0.729 |
|  | (0.951-0.980) | (0.622-0.819) |
| LVEF ≤ 50% | 0.963 | 0.903 |
|  | (0.948-0.982) | (0.867-0.935) |
| **Age** |  |  |
| age ≥ 65  (n=263) | 0.959 | 0.868 |
|  | (0.936-0.982) | (0.806-0.925) |
| age < 65  (n=417) | 0.974 | 0.918 |
|  | (0.939-1.00) | (0.852-0.970) |
| **Sex** |  |  |
| Male  (n=350) | 0.961 | 0.890 |
|  | (0.933-0.990) | (0.838-0.937) |
| Female  (n=331) | 0.984 | 0.866 |
|  | (0.972-0.996) | (0.779-0.938) |
| **BMI** |  |  |
| BMI < 25  (n=396) | 0.965 | 0.891 |
|  | (0.940-0.991) | (0.839-0.935) |
| 25 ≤ BMI ≤ 30  (n=227) | 0.980 | 0.837 |
|  | (0.965-0.995) | (0.724-0.930) |
| BMI > 30  (n=58) | 0.993 | 0.963 |
|  | (0.978-1.00) | (0.885-1.00) |
| **HTN** |  |  |
| HTN  (n=288) | 0.967 | 0.863 |
|  | (0.947-0.987) | (0.794-0.922) |
| non-HTN  (n=393) | 0.972 | 0.907 |
|  | (0.941-1.00) | (0.840-0.957) |
| **DM** |  |  |
| DM  (n=141) | 0.943 | 0.914 |
|  | (0.901-0.985) | (0.861-0.958) |
| non-DM  (n=540) | 0.983 | 0.865 |
|  | (0.971-0.994) | (0.791-0.922) |
| **CAD** |  |  |
| CAD  (n=104) | 0.930 | 0.879 |
|  | (0.882-0.977) | (0.797-0.944) |
| non-CAD  (n=577) | 0.975 | 0.888 |
|  | (0.950-1.00) | (0.827-0.939) |
| **CKD** |  |  |
| CKD  (n=31) | 0.933 | 0.934 |
|  | (0.850-1.00) | (0.844-0.991) |
| non-CKD  (n=650) | 0.971 | 0.875 |
|  | (0.951-0.991) | (0.820-0.921) |
| **SR** |  |  |
| NSR  (n=613) | 0.972 | 0.877 |
|  | (0.950-0.995) | (0.824-0.292) |
| non-NSR  (n=68) | 0.915 | 0.893 |
|  | (0.851-0.979) | (0.808-0.958) |
| **AF** |  |  |
| AF  (n=43) | 0.876 | 0.865 |
|  | (0.775-0.977) | (0.756-0.956) |
| non-AF  (n=638) | 0.973 | 0.886 |
|  | (0.953-0.994) | (0.836-0.932) |
| **PM** |  |  |
| PM  (n=10) | 1.00 | 1.00 |
|  | (1.00-1.00) | (1.00-1.00) |
| non-PM  (n=671) | 0.970 | 0.875 |
|  | (0.952-0.989) | (0.823-0.919) |
| **LVH** |  |  |
| LVH  (n=66) | 0.976 | 0.947 |
|  | (0.948-1.00) | (0.883-0.990) |
| non-LVH  (n=615) | 0.969 | 0.868 |
|  | (0.947-0.991) | (0.812-0.914) |
| **BBB** |  |  |
| BBB  (n=37) | 0.960 | 0.943 |
|  | (0.907-1.00) | (0.853-0.993) |
| non-BBB  (n=644) | 0.971 | 0.873 |
|  | (0.950-0.991) | (0.825-0.920) |
| **LOW** |  |  |
| LOW  (n=16) | 0.945 | 0.877 |
|  | (0.830-1.00) | (0.600-1.00) |
| non-LOW  (n=665) | 0.971 | 0.881 |
|  | (0.953-0.990) | (0.837-0.921) |

BMI, body mass index; HTN, hypertension; DM, diabetes mellitus; CAD, coronary artery disease; CKD, chronic kidney disease; SR, sinus rhythm; AF, atrial fibrillation; PM, pacemaker; LVH, left ventricular hypertrophy; BBB, right and left bundle branch block; LOW, low voltage

**Supplemental Table 4.** Performance comparison between NT-proBNP and AiTiALVSD among 96 patients

|  | AUROC (95% CI) | Sensitivity (95% CI) | Specificity (95% CI) | PPV (95% CI) | NPV (95% CI) |
| --- | --- | --- | --- | --- | --- |
| NT-proBNP | 0.72 | 0.889 | 0.551 | 0.436 | 0.927 |
|  | (0.635-0.804) | (0.770-1.0) | (0.433-0.668) | (0.305-0.567) | (0.847-1.0) |
| AiTiALVSD | 0.905 | 0.926 | 0.884 | 0.758 | 0.968 |
|  | (0.842-0.968) | (0.827-1.0) | (0.809-0.960) | (0.611-0.904) | (0.925-1.0) |

AUROC, area under the receiver operating characteristic curve; PPV, positive predictive value; NPV, negative predictive value; CI, confidence interval

**Supplemental Table 5.** Phenotyping of AiTiALVSD-positive LVSD cases

|  | **Phenotype A** | | **Phenotype B** | **Phenotype C** | **Phenotype D** | **Phenotype E** | **Total** |
| --- | --- | --- | --- | --- | --- | --- | --- |
|  | (n=16) | | (n=13) | (n=32) | (n=49) | (n=13) | (n=123) |
| Age, year | 66.25 ± 11.87 | | 69.77 ± 11.30 | 69.22 ± 14.37 | 69.90 ± 14.06 | 68.15 ± 14.06 | 69.05 ± 13.45 |
| Male, n (%) | 13 (81.25) | | 9 (69.23) | 20 (62.5) | 34 (69.39) | 7 (53.85) | 83 (67.48) |
| Height, cm | 166.19 ± 8.00 | | 165.22 ± 8.42 | 162.86 ± 10.08 | 163.86 ± 9.95 | 164.17 ± 12.01 | 164.08 ± 9.74 |
| Weight, kg | 71.79 ± 16.42 | | 74.85 ± 13.13 | 62.52 ± 11.84 | 66.82 ± 15.70 | 67.81 ± 19.56 | 67.30 ± 15.34 |
| **Medical History** | | | | | | | |
| HTN, n (%) | 9 (56.25) | | 9 (69.23) | 13 (40.63) | 26 (53.06) | 5 (38.46) | 62 (50.41) |
| DM, n (%) | 9 (56.25) | | 8 (61.54) | 14 (43.75) | 19 (38.78) | 4 (30.77) | 54 (43.9) |
| CAD, n (%) | | 11 (68.75) | 3 (23.08) | 10 (31.25) | 23 (46.94) | 1 (7.69) | 48 (39.02) |
| CKD, n (%) | | 4 (25) | 1 (7.69) | 8 (25) | 7 (14.29) | 1 (7.69) | 21 (17.07) |
| **ECG feature** | | | | | | | |
| HR, bpm | | 75.50 ± 11.26 | 88.00 ± 12.45 | 74.88 ± 12.70 | 67.65 ± 9.86 | 101.08 ± 18.47 | 76.24 ± 15.95 |
| PR interval, ms | | 181.62 ± 30.09 | 192.86 ± 21.57 | 177.33 ± 37.00 | 182.12 ± 45.31 | 170.60 ± 27.01 | 180.99 ± 38.28 |
| QT interval, ms | | 421.38 ± 38.45 | 397.69 ± 39.58 | 427.09 ± 36.59 | 422.02 ± 40.80 | 373.38 ± 42.77 | 415.54 ± 42.31 |
| QRS duration, ms | | 112.38 ± 20.66 | 122.62 ± 31.94 | 119.78 ± 25.13 | 111.80 ± 26.70 | 108.23 ± 30.50 | 114.72 ± 26.59 |
| QTc interval, ms | | 469.56 ± 32.62 | 477.92 ± 24.92 | 473.81 ± 38.85 | 445.37 ± 39.42 | 479.08 ± 35.57 | 462.92 ± 39.03 |
| P axis | | 53.44 ± 20.28 | 54.14 ± 15.81 | 58.05 ± 23.01 | 43.56 ± 32.23 | 39.00 ± 67.46 | 49.36 ± 30.28 |
| R axis | | 41.62 ± 32.86 | 17.00 ± 48.85 | 22.50 ± 64.52 | 19.29 ± 59.53 | 17.69 ± 63.84 | 22.62 ± 57.26 |
| T axis | | 97.87 ± 50.35 | 57.08 ± 69.09 | 77.00 ± 103.26 | 90.59 ± 82.26 | 39.23 ± 36.59 | 78.91 ± 81.05 |
| **ECG LVSD feature** | | | | | | | |
| Prolonged QRS | | 8 (50) | 5 (38.46) | 16 (50) | 14 (28.57) | 3 (23.08) | 46 (37.40) |
| LAD | | 2 (12.50) | 4 (30.77) | 8 (25) | 13 (26.53) | 4 (30.77) | 31 (25.20) |
| RAD | | 1 (6.25) | 0 (0) | 4 (12.50) | 7 (14.29) | 2 (15.38) | 14 (11.38) |
| QT prolongation | | 11 (68.75) | 11 (84.62) | 23 (71.88) | 16 (32.65) | 10 (76.92) | 71 (57.72) |
| Abnormal QRS-T axis | | 0 (0) | 1 (7.69) | 5 (15.62) | 5 (10.20) | 0 (0) | 11 (8.94) |
| delayed ID | | 9 (56.25) | 5 (38.46) | 12 (37.50) | 23 (46.94) | 3 (23.08) | 52 (42.28) |
| Abnormal Q | |  |  |  |  |  |  |
| *V2, V3, V4* | | 4 (25.00) | 0 (0) | 1 (3.12) | 10 (20.41) | 0 (0) | 15 (12.20) |
| *V1, V2* | | 5 (31.25) | 0 (0) | 2 (6.25) | 12 (24.49) | 0 (0) | 19 (15.45) |
| *V5, V6, I, aVL* | | 1 (6.25) | 0 (0) | 2 (6.25) | 3 (6.12) | 0 (0) | 6 (4.88) |
| *II, aVF, III* | | 1 (6.25) | 0 (0) | 4 (12.50) | 7 (14.29) | 1 (7.69) | 13 (10.57) |
| T wave inversion | |  |  |  |  |  |  |
| *I, V5, or V6* | | 5 (31.25) | 0 (0) | 7 (21.88) | 7 (14.29) | 1 (7.69) | 20 (16.26) |
| *V2-4* | | 5 (31.25) | 0 (0) | 2 (6.25) | 4 (8.16) | 2 (15.38) | 13 (10.57) |
| *II, aVF* | | 0 (0) | 0 (0) | 4 (12.50) | 1 (2.04) | 0 (0) | 5 (4.07) |
| ST elevation | |  |  |  |  |  |  |
| *V2-4* | | 3 (18.75) | 4 (30.77) | 3 (9.38) | 8 (16.33) | 1 (7.69) | 19 (15.45) |
| *V1-2* | | 0 (0) | 2 (15.38) | 2 (6.25) | 4 (8.16) | 1 (7.69) | 9 (7.32) |
| *V5, V6, I, aVL* | | 0 (0) | 0 (0) | 0 (0) | 0 (0) | 0 (0) | 0 (0) |
| *II, III, aVF* | | 0 (0) | 0 (0) | 0 (0) | 0 (0) | 0 (0) | 0 (0) |
| ST depression | |  |  |  |  |  |  |
| *V2-4* | | 0 (0) | 0 (0) | 3 (9.38) | 0 (0) | 1 (7.69) | 4 (3.25) |
| *V1-2* | | 0 (0) | 0 (0) | 0 (0) | 0 (0) | 1 (7.69) | 1 (0.81) |
| *V5, V6, I, aVL* | | 0 (0) | 0 (0) | 8 (25.00) | 0 (0) | 1 (7.69) | 9 (7.32) |
| *II, III, aVF* | | 0 (0) | 0 (0) | 1 (3.12) | 1 (2.04) | 0 (0) | 2 (1.63) |
| Tall T | |  |  |  |  |  |  |
| *V2-4* | | 1 (6.25) | 3 (23.08) | 2 (6.25) | 1 (2.04) | 1 (7.69) | 8 (6.50) |
| *V1-2* | | 0 (0) | 2 (15.38) | 1 (3.12) | 0 (0) | 1 (7.69) | 4 (3.25) |
| *V5, V6, I, aVL* | | 0 (0) | 0 (0) | 0 (0) | 0 (0) | 0 (0) | 0 (0) |
| *II, III, aVF* | | 0 (0) | 0 (0) | 0 (0) | 1 (2.04) | 0 (0) | 1 (0.81) |
| **ECG device interpretation** | | | | | | | |
| SR | | 16 (100) | 8 (61.54) | 22 (68.75) | 34 (69.39) | 5 (38.46) | 85 (69.11) |
| LVH | | 2 (12.5) | 2 (15.38) | 12 (37.50) | 6 (12.24) | 5 (38.46) | 27 (21.95) |
| AF | | 0 (0) | 5 (38.46) | 7 (21.88) | 7 (14.29) | 7 (53.85) | 26 (21.14) |
| LBBB | | 1 (6.25) | 2 (15.38) | 1 (3.12) | 4 (8.16) | 1 (7.69) | 9 (7.32) |
| RBBB | | 1 (6.25) | 1 (7.69) | 3 (9.38) | 0 (0) | 2 (15.38) | 7 (5.69) |
| PM | | 0 (0) | 0 (0) | 2 (6.25) | 4 (8.16) | 0 (0) | 6 (4.88) |
| **Echocardiographic findings** | | | | | | | |
| EF, % | | 36.75 ± 8.55 | 41.46 ± 13.22 | 35.97 ± 11.41 | 39.37 ± 11.05 | 38.19 ± 11.55 | 38.24 ± 11.12 |
| LVIDs, mm | | 42.90 ± 10.20 | 45.23 ± 13.53 | 48.76 ±. 0.77 | 44.48 ± 9.90 | 43.36 ± 12.00 | 45.46 ± 10.88 |
| LVIDd, mm | | 59.20 ± 8.64 | 58.31 ± 11.02 | 60.63 ± 8.64 | 57.80 ± 8.66 | 53.40 ± 9.29 | 58.33 ± 9.09 |
| Mitral inflow E cm/s | | 63.25 ± 26.09 | 87.17 ± 17.72 | 86.52 ± 37.22 | 68.40 ± 21.12 | 67.25 ± 22.58 | 75.01 ± 27.40 |
| Mitral inflow A cm/s | | 68.38 ± 22.60 | 83.67 ± 15.59 | 67.64 ± 22.84 | 72.83 ± 24.68 | 70.67 ± 30.66 | 71.86 ± 23.19 |
| Deceleration time, ms | | 210.88 ± 53.98 | 201.82 ± 83.70 | 204.93 ± 83.34 | 217.89 ± 56.21 | 150.88 ± 58.94 | 205.34 ± 69.03 |
| Septal E′ cm/s | | 4.54 ± 1.85 | 11.58 ± 20.46 | 4.23 ± 1.88 | 4.97 ± 1.60 | 5.65 ± 2.02 | 5.76 ± 7.99 |
| Septal A′ cm/s | | 7.41 ± 2.17 | 6.83 ± 2.14 | 5.88 ±. .83 | 6.95 ± 2.10 | 6.33 ± 2.52 | 6.69 ± 2.32 |
| E/E′ ratio | | 17.51 ± 12.83 | 18.34 ± 12.85 | 21.71 ± 9.69 | 15.52 ± 8.27 | 12.59 ± 3.39 | 17.38 ± 9.79 |
| E/A ratio | | 1.27 ± 1.41 | 1.04 ± 0.21 | 1.40 ± 0.50 | 1.10 ± 0.77 | 0.87 ± 0.42 | 1.18 ± 0.90 |
| PASP, mmHg | | 31.50 ± 14.65 | 29.15 ± 8.15 | 30.96 ± 13.92 | 26.18 ± 8.86 | 23.80 ± 4.71 | 28.17 ± 10.89 |
| **LVSD score and reference standard** | | | | | | | |
| AiTiALVSD score | | 42.98 ± 23.20 | 33.24 ± 25.71 | 55.87 ± 28.50 | 40.01 ± 22.43 | 37.75 ± 26.41 | 43.57 ± 25.77 |
| LVSD (True positive) | | 13 (81.25) | 8 (61.54) | 24 (75) | 33 (67.35) | 10 (76.92) | 88 (71.54) |
| non-LVSD (False positive | | 3 (18.75) | 5 (38.46) | 8 (25) | 16 (32.65) | 3 (23.08) | 35 (28.46) |
| Values are expressed as n(%), mean ± standard deviation; HTN, hypertension; DM, diabetes mellitus; CAD, coronary artery disease; CKD, chronic kidney disease; SR, sinus rhythm; AF, atrial fibrillation; PM, pacemaker; LVH, left ventricular hypertrophy; BBB, right and left bundle branch block | | | | | | | |

**Supplemental Text**

**Supplemental Text 1. AiTiALVSD algorithm development**

We developed a model based on a residual neural network (ResNet) utilizing the PyTorch library and Python for programming. ResNet, typically used for image processing, works by utilizing convolutions to identify intricate patterns within datasets.^34^ ECG classifications possess complex characteristics for classification tasks in image and time series domains. Such tasks require extracting rhythm and morphological features from ECG. Figure S1 shows a structure of our model. We employed architectures with a stem block, 12 residual blocks, and a single fully connected network to detect patterns in these features. Each block extracts the features of the ECG and passes them to the next block. We defined a feature block as a group of three residual blocks where AiTiALVSD contains four feature blocks. The residual block consists of a set of layers: a one-dimensional convolutional neural network, batch normalization, rectified linear unit (ReLU) activations, another one-dimensional convolutional neural network, additional batch normalization, a subsequent ReLU activation, a dropout layer and skip connection. The stem block contains a single layer with a skip connection and max pooling.

Our ECG data were collected from three hospitals in the Republic of Korea, excluding the hospital utilized in this study. All digitalized 12-lead ECGs were measured with a sampling rate of 500 Hz for 10 seconds. To facilitate computations during the training phase, we represented the ECGs as a 12x5000 matrix and down sampled them. Additionally, we employed data augmentation to enhance the model's generalization.^35^ Our dataset was split into three parts, with 80% of the data used for training, 15% for validation during training, and 5% for testing the final model. ECGs from a single patient were not split. The positive label was defined as an ejection fraction (EF) of 40% or less as observed on cardiac ultrasound by the diagnostic and treatment guidelines for heart failure presented by the European Society of Cardiology.^1^ Conversely, the zero labels indicated an EF of greater than 40%.

During training, we used the Adam optimizer with the cosine warm-up optimization scheduler to update the model weights and the focal loss function.^36^ We employed the population-based training scheduler to optimize various hyperparameters, including learning rate, weight decay in the optimizer, and network parameters such as kernel size, number of layers, blocks, and dropout rates.^37^ This scheduler was run for 150 epochs to evaluate different hyperparameters and identify the optimal model architecture based on a combined value of AUROC, AUPRC, loss, and F1-score performance on the validation set.

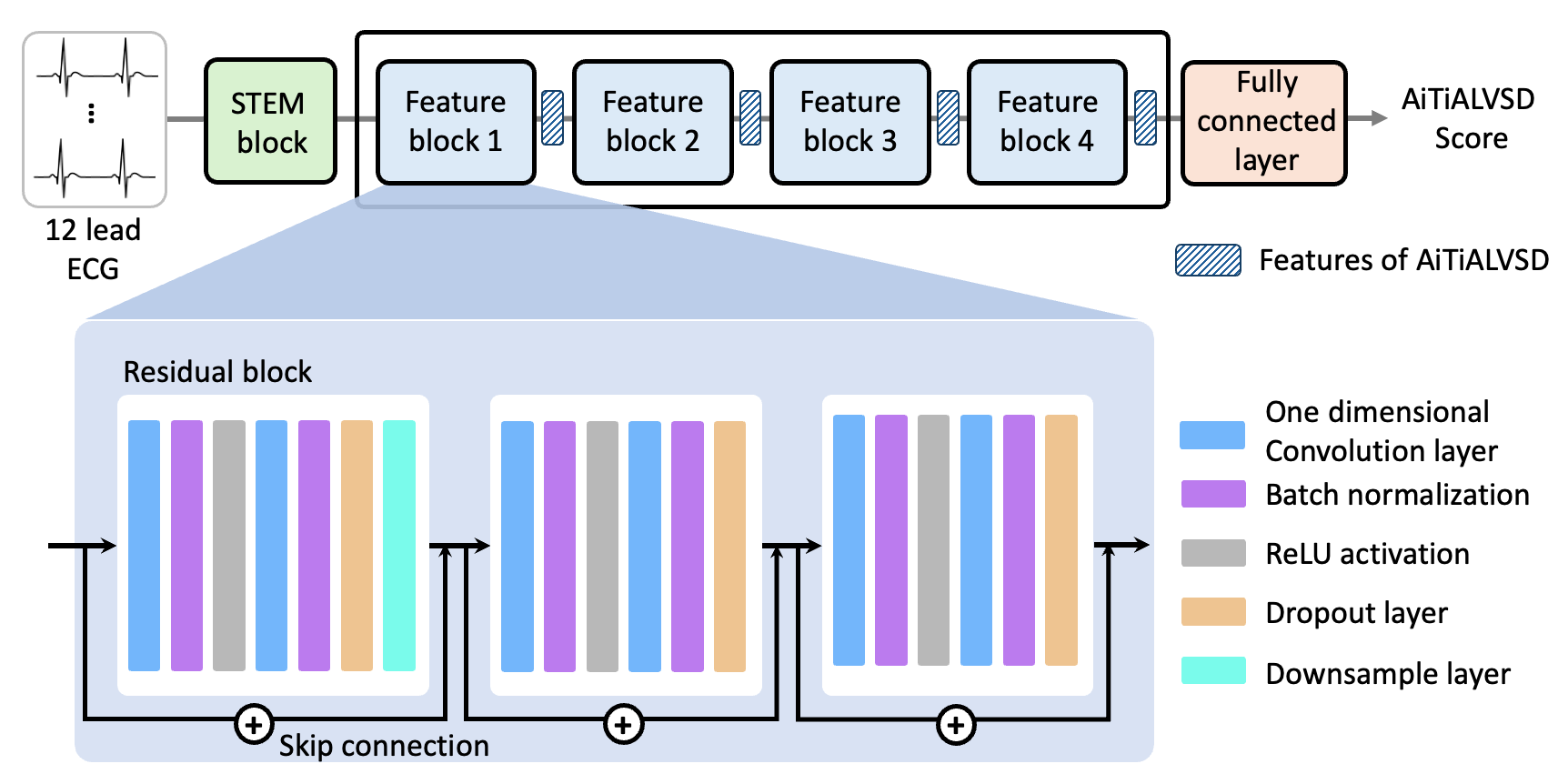

**Figure S1. Structure of AiTiALVSD.** It shows structure of AiTiALVSD and process to compute an AiTiALVSD score. The process begins with a 12-lead ECG input which goes through a STEM block, followed by four sequential feature blocks, and finally culminates in a fully connected layer that outputs the AITIALVSD score. Each feature block contains three residual blocks with skip connections. Inside the residual blocks, there are layers represented by colored bars, each color corresponding to a different type of operation: one-dimensional convolution layers, batch normalization, ReLU activation functions, dropout layers, and downsample layer.

**Supplemental Text 2. Model Card of AiTiALVSD for model development**

Table S2.1 shows demographic distribution of dataset for AiTiALVSD development. This dataset consists of 353,845 records, divided into 291,564 training entries (79.9% of the total), 55,320 validation entries (15.1%), and 17,961 testing entries (5.0%).

**Table S2.1 Data distribution of AiTiALVSD development**

|  |  | **Train**  **dataset** | | **Validation**  **dataset** | | **Test**  **dataset** | | **Total** |
| --- | --- | --- | --- | --- | --- | --- | --- | --- |
|  | **# of ECG record** | 291564 | | 55320 | | 17961 | | 364845 |
|  | **Hospitals** | | | | | | | |
|  | **Hospital A** | 193702 | | 37074 | | 11883 | | 242659 |
|  | **Hospital B** | 60622 | | 11353 | | 3737 | | 75712 |
|  | **Hospital C** | 22053 | | 4019 | | 1378 | | 27450 |
|  | **Hospital D** | 15187 | | 2874 | | 963 | | 19024 |
| **Gender** | | | | | | | | |
|  | **Female** | 124277 | 23333 | | 7688 | | 155298 | |
|  | **Male** | 162139 | 30945 | | 9954 | | 203038 | |
|  | **Unknown** | 5148 | 1042 | | 319 | | 6509 | |
| **Age group** | | | | | | | | |
|  |  | 69 | 14 | | 4 | | 87 | |
|  | **~20** | 1683 | 294 | | 97 | | 2074 | |
|  | **20~30** | 7828 | 1393 | | 555 | | 9776 | |
|  | **30~40** | 14944 | 2854 | | 1002 | | 18800 | |
|  | **40~50** | 32415 | 6167 | | 1937 | | 40519 | |
|  | **50~60** | 57633 | 10552 | | 3934 | | 72119 | |
|  | **60~70** | 66647 | 12491 | | 3926 | | 83064 | |
|  | **70~80** | 69352 | 13345 | | 4047 | | 86744 | |
|  | **80~90** | 31864 | 6351 | | 1968 | | 40183 | |
|  | **90~100** | 3045 | 636 | | 132 | | 3813 | |
|  | **100~** | 44 | 4 | | 0 | | 48 | |
|  |  | 0 | 1 | | 0 | | 1 | |
|  |  | 1 | 0 | | 0 | | 1 | |

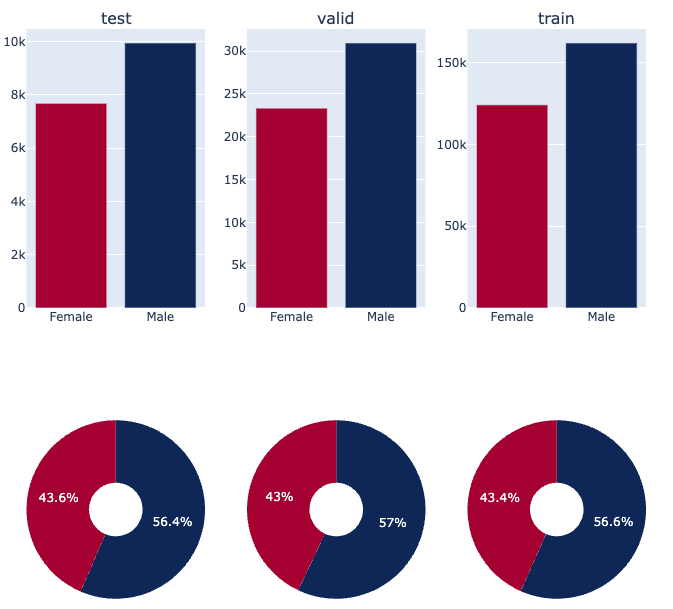

**Figure S2.1** Proportion of gender in datasets.

Gender distribution within the dataset is detailed as follows: females account for 124,277 records in the training set (43.6%), 23,333 in validation (43.0%), and 7,688 in testing (43.4%). Males are represented by 162,139 records in the training set (56.4%), 30,945 in validation (57.0%), and 9,954 in testing (56.6%). Unknown gender records comprise 5,148 in training, 1,042 in validation, and 319 in testing.

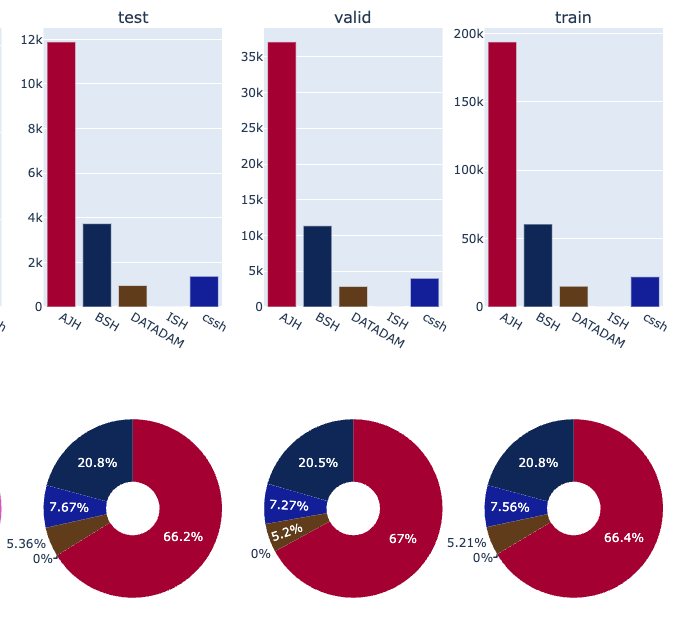

**Figure S2.2** Proportion of hospital in each dataset.

Hospital affiliation data is categorized across four hospitals: Hospital A, B, C, and D. Hospital A is the most represented with 193,702 records in the training set (66.2%), 37,074 in validation (67.0%), and 11,883 in testing (66.4%). Hospital B contributes 60,622 records to training (20.8%), 11,353 to validation (20.5%), and 3,737 to testing (20.8%). Hospital C and D provide smaller but significant contributions, ensuring a diverse range of medical practices and patient demographics are represented, enhancing the dataset's applicability to various research contexts.

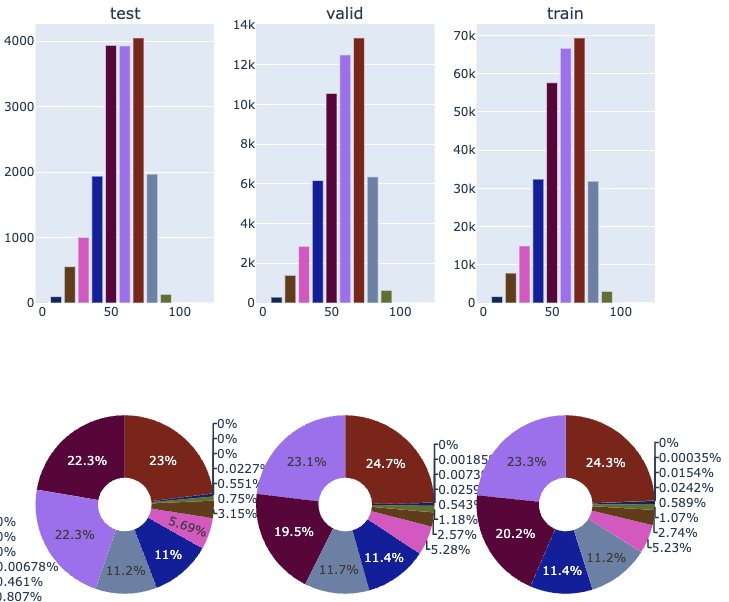

**Figure S2.2** Proportion of age group in each dataset.

The age group distribution is notably skewed towards older individuals, particularly beneficial for research on age-related conditions. The most represented age groups are 50-80 years, with the 70-80 age group alone accounting for 4,047 records in training (1.4%), 13,345 in validation (24.1%), and 69,352 in testing (386.3%).

In summary, the dataset's composition—characterized by a balanced gender distribution, diverse hospital affiliations, and a focus on broad age groups—offers a solid foundation for a wide range of medical research topics. The proportional representation of different demographics and affiliations within the dataset underscores its potential for generating nuanced and broadly applicable health insights.

The dataset's structure and diversity are conducive to a wide array of medical research topics, from gender and age-related studies to analyses based on hospital-specific data. Its comprehensive nature allows for the development of nuanced and broadly applicable health insights.

**Supplemental Text 3. Interpretation methods for model transparency**

**A. Concept-based Method: Test concept activation vector (TCAV)**

TCAV is a technique for interpreting the decisions made by a deep neural network. It is designed to provide insights into the internal workings of a model in a way that is understandable to humans.

The main idea behind TCAV is to represent high-level concepts as vectors in the activation space of a neural network layer. These vectors, known as concept activation vectors (CAVs), can be used to measure the extent to which a given concept is utilized by the model for prediction. By calculating a TCAV score for a concept, we can quantify the overall importance of that concept for the decisions made by the model. Figure S3.1 shows a process of TCAV. Below is the detailed process of TCAV we applied.

1. **Concept Identification:** The first step in TCAV involves identifying the concepts of interest. These concepts can be anything that you believe might have an influence on the decisions made by the model. For example, in an image classification task, concepts could be colors, textures, shapes, or even specific objects. We utilized the Physionet open dataset to define concepts and construct a concept dataset. The Physionet challenge dataset is an integrated dataset of ECGs collected from six different hospitals. The Physionet dataset comprises a total of N ECGs, each labeled with one of 26 arrhythmias, making it a multilabel ECG dataset. Each ECG possesses one or more arrhythmia labels. Figure S3.1 displays the label distribution of the Physionet dataset.
   We defined nine target concepts using these labels. Our objective was to ascertain whether AiTiALVSD positively utilizes these target concepts in determining LVSD by using the TCAV score. The nine concepts were as follows: atrial fibrillation and atrial flutter (AF), left bundle branch block (LBBB), right bundle branch block (RBBB), left axis deviation (LAD), right axis deviation (RAD), prolonged QT, nonspecific intraventricular conduction disorder (conduction disorder), and combined abnormal q wave and abnormal t wave (abnormal Q, T wave). The AF and abnormal Q wave concepts were defined by combining two labels. The AF concept included both atrial fibrillation and atrial flutter. The abnormal Q wave concept included both abnormal T wave and T wave inversion labels.
   In constructing the concept datasets, we ensured that there were no overlapping ECGs among them by extracting distinct ECGs. From the remaining labels not used for the concepts, we randomly selected approximately 200 samples to create 10 random concept datasets. When constructing a concept dataset, we ensured that the number of data extracted from each data source was balanced. This was to prevent any specific concept dataset from having a bias toward a particular data source. If a data source was lacking labels, we supplemented it with additional data from another source. Table S1 shows the distribution of data sources in each concept dataset. As a result, we established nine target concept datasets and 10 random concept datasets.
2. **Feature Extraction:** Once the concepts are identified, the next step is to extract hidden features of each block. Figure S3.1 (2) shows an example of hidden feature extraction from the first block of AiTiALVSD. Following a feature representation for AiTiALVSD to predict LVSD, the features of all samples from the concept datasets are extracted.
3. **Concept Activation Vector (CAV):** A Concept Activation Vector is a direction in the activation space of a neural network layer that represents a concept. To create a CAV, a linear classifier is trained (such as a logistic regression or SVM) to distinguish between the above two sets of targets that can random concept features. The vector orthogonal to the decision boundary of this classifier is defined as the CAV for the target concept.
4. **Extraction feature from positive LVSD samples:** features of 300 positive LVSD samples are extracted using the same process as step (2).
5. **Testing with CAVs (TCAV Score Calculation):** After the CAV and positive LVSD features are created, they can be used to interpret the decisions made by the model. For given LVSD features, it moves slightly onto the CAV. We check whether the moved LVSD feature of the AiTiALVSD score increases when predicted again. By repeating this process for all LVSD samples and averaging the results, a TCAV score for the target concept is calculated as follows: This score quantifies the overall importance of the concept for the decisions made by the model.

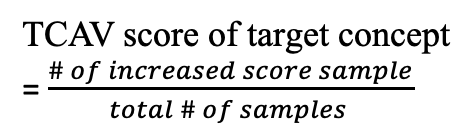

6. **Interpretation:** If a concept has a high TCAV score, it means that the model places much positive importance on that concept when making decisions. If a concept has a low TCAV score, it means that the concept is not important for the model's decisions.

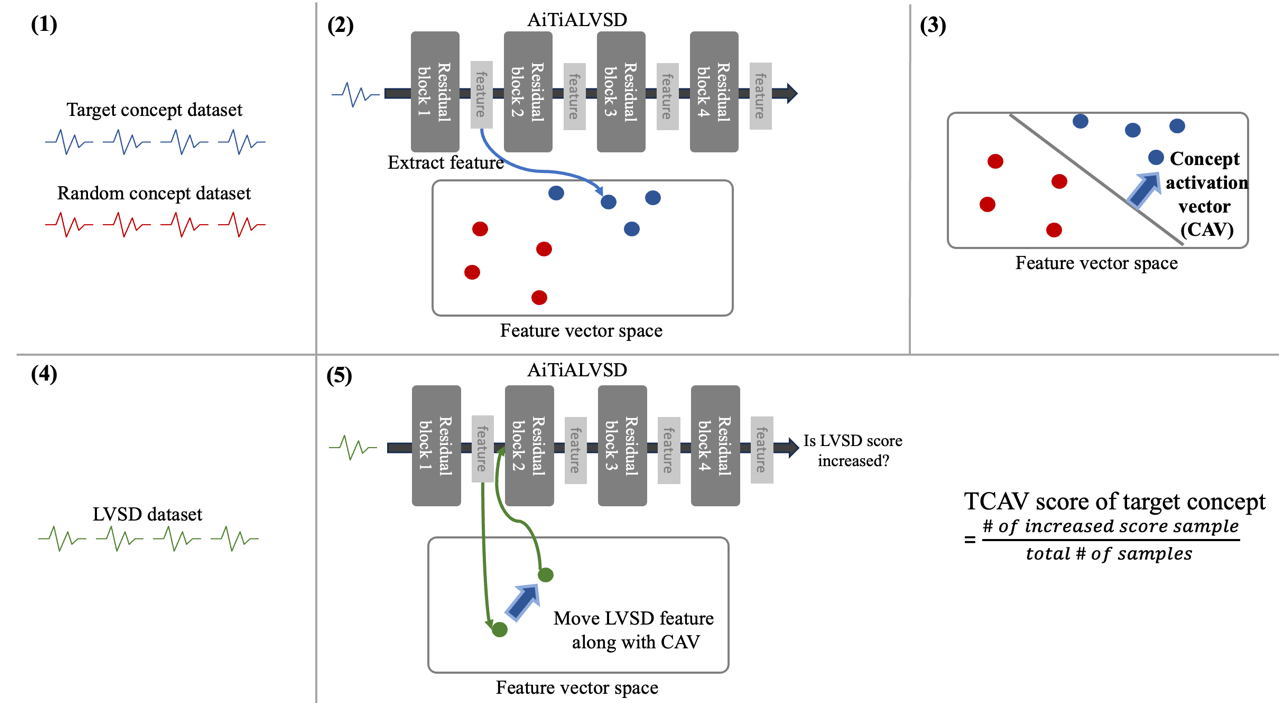

**Figure S3.1** Process of TCAV

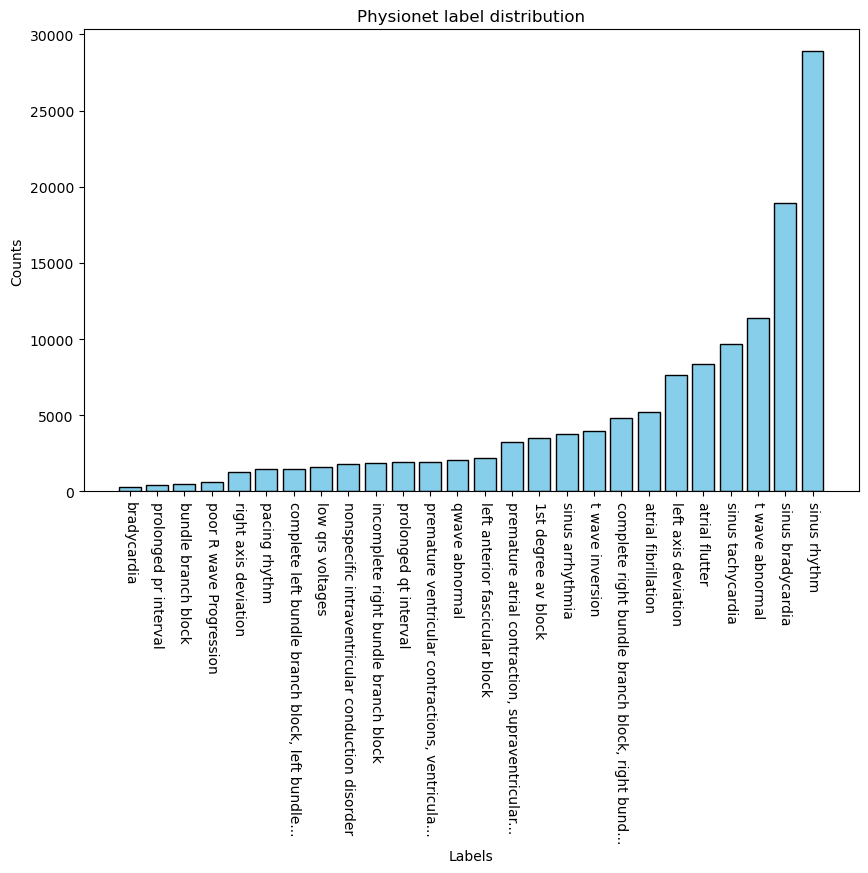

**Figure S3.2** Histogram of label count in the Physionet dataset

**Table S3.1** Distribution of data sources for each concept dataset

|  | **Ningbo** | **Georgia** | **Chapman Shaoxing** | **PTBXL** | **CPSC2018** | **CPSC2018 extra** |
| --- | --- | --- | --- | --- | --- | --- |
| **right axis deviation** | 79 | 40 | 40 | 40 | - | 1 |
| **complete left bundle branch block, left bundle...** | 33 | 33 | 33 | 63 | 33 | 5 |
| **nonspecific intraventricular conduction disorder** | 40 | 40 | 40 | 76 | - | 4 |
| **prolonged QT interval** | 40 | 76 | 40 | 40 | - | 4 |
| **Q wave abnormal** | 79 | 40 | 40 | 40 | - | 1.0 |
| **complete right bundle branch block, right bund...** | 33 | 33 | 33 | 33 | 35 | 33 |
| **left axis deviation** | 50 | 50 | 50 | 50 | - | - |
| **atrial fibrillation+atrial flutter** | 35 | 33 | 33 | 33 | 33 | 33 |
| **T wave abnormal + T wave inversion** | 54 | 40 | 40 | 40 | - | 26 |
| **random_concept_0** | 33 | 33 | 33 | 33 | 33 | 33 |
| **random_concept_1** | 33 | 33 | 33 | 33 | 33 | 33 |
| **random_concept_2** | 33 | 33 | 33 | 33 | 33 | 33 |
| **random_concept_3** | 33 | 33 | 33 | 33 | 33 | 33 |
| **random_concept_4** | 33 | 33 | 33 | 33 | 33 | 33 |
| **random_concept_5** | 33 | 33 | 33 | 33 | 33 | 33 |
| **random_concept_6** | 33 | 33 | 33 | 33 | 33 | 33 |
| **random_concept_7** | 33 | 33 | 33 | 33 | 33 | 33 |
| **random_concept_8** | 33 | 33 | 33 | 33 | 33 | 33 |
| **random_concept_9** | 33 | 33 | 33 | 33 | 33 | 33 |

Ningbo, Ningbo Database; Georgia, The Georgia 12-lead ECG Challenge (G12EC) Database; PTB, PTB and PTB-XL Database

**Supplemental Text 4. Sample size calculation**

In the prior studies that developed the software, the respective AUROCs were ranging from 0.877 to 0.9790.925 and 0.963; however, considering the possibility of overestimation in this study.^7,8^ We assumed an expected AUROC of 0.85 for the strict clinical trial, accounting for the potential for limited performance. Using the AUROC values of BNP or NT-proBNP, the conventional representative indicators for screening LVSD, as the basis for determining the minimum clinical performance standard, the reported performance levels range widely from 0.56 to 0.90.^38,39^ Consequently, we set the validity evaluation criteria for this clinical trial at AUROC 0.75 and determined a superiority margin of 0.1 to calculate the sample size. Furthermore, since the prevalence of LVSD varies among communities and hospitals, we set the prevalence at 15%.^5,7,40^ Sample size calculation was performed using PASS 15 (NCSS, LLC. Kaysville, USA) software. The performance estimate of AiTiALVSD was set at AUROC 0.85, the clinical validity evaluation criterion at AUROC 0.75, and the significance level (α) and statistical power (1-β) at 0.05 and 95%, respectively. Considering a dropout rate of 5%, the total sample size was 688 participants (103 LVSD and 585 Non-LVSD).

**B. Cluster Analysis**

We performed clustering analysis to analyze the characteristics of LVSD that AiTiALVSD recognizes. By comparing the ECG features of each cluster group and visually verifying them, we aimed to distinguish specific ECG differences. The clustering algorithm was executed using the sklearn python package version 1.1.1. The details of the clustering analysis are as follows:

**1. Selection of ECG samples and AiTiALVSD features:** To understand the patterns of ECGs positively predicted by AiTiALVSD, we only used positively predicted ECG samples. As a result, the cluster algorithm was trained on a total of 60,125 positively predicted ECG samples in the development dataset, and the clustering results were visualized using positive predicted samples from the clinical trial. We utilized features extracted from the first block of AiTiALVSD as input for clustering. Features from the first block consisted of 64 channels, each with a size of 313. To address the curse of dimensionality and computational resource issues, we averaged each channelwise aggregation, converting them into 64-sized features. We trained the cluster algorithm using these transformed 64-sized features.

**2. Clustering algorithm and determining the number of clusters:** For the cluster analysis, we employed the K-means algorithm. To determine the optimal number of clusters, we used the elbow method. We set the number of clusters to five, which is most appropriate to capture the inherent patterns within the data.

**3. Result visualization:** To visualize the results of clustering, we employed the t-distributed stochastic neighbor embedding (T-SNE) method for dimensionality reduction. Using T-SNE, we reduced the 64-sized features into 2D data and visualized the results through a scatter plot. Supplemental Figure S3.3-4 shows the clustering visualization results for both the training dataset and the clinical dataset. We observed distinct clusters in the clinical trial dataset, which indicates that the AiTiALVSD features internal phenotypes based on ECG characteristics.

**4. Selection of representative ECGs:** We assumed that each cluster defines a phenotype and compared the baseline characteristics of each cluster using a t test. Furthermore, to select a representative ECG, we chose the ECG closest to the tcentroid of each cluster from the clinical trial dataset. Figures S3.3 and S3.4 display the phenotype ECG selected from each cluster in the training and clinical trial datasets.

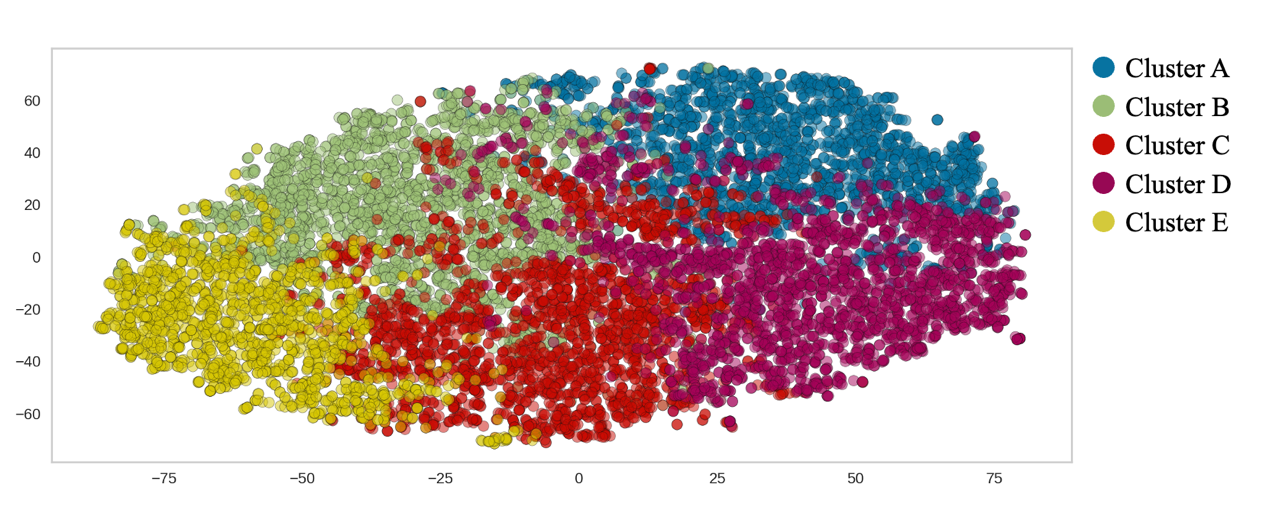

**Figure S3.3** Clustering visualization using T-SNE in the development dataset

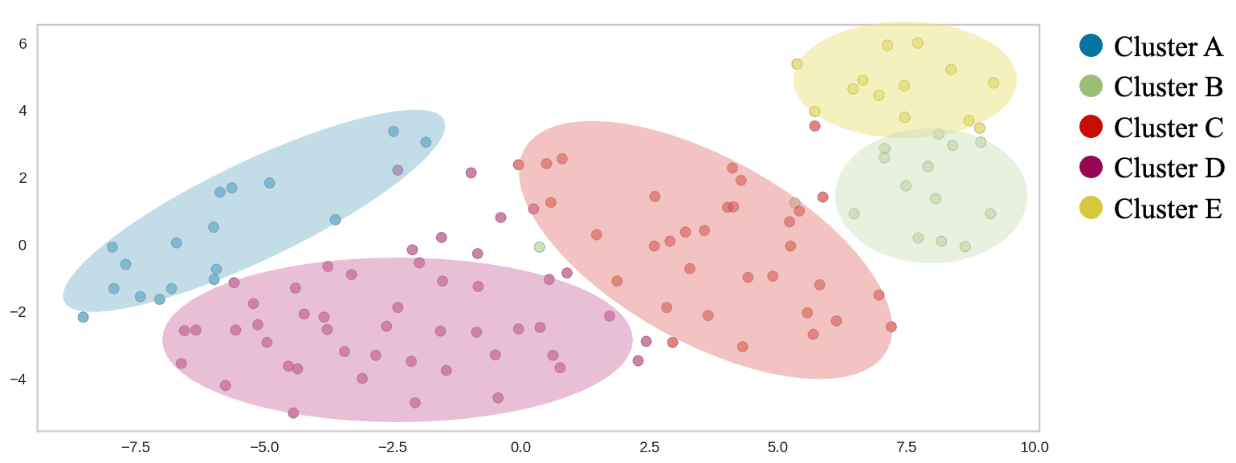

**Figure S3.4** Clustering visualization using T-SNE in the clinical trial dataset
